# Expanding the clinical spectrum of autoimmune inflammatory myopathies with prominent B cell aggregates: a case series

**DOI:** 10.64898/2026.03.01.26347357

**Authors:** Barath Ramanathan, Hao Cheng Shen, Marie Hudson, Yves Troyanov, Océane Landon-Cardinal, Geneviève Gyger, Erin O’Ferrall, Benjamin Ellezam, Jason Karamchandani, Celia del Carmen Crespo Oliva, Zoé Gerber, Dominique Jean, Marilyne Labrie, Valérie Leclair, Hugues Allard-Chamard

## Abstract

Autoimmune inflammatory myopathies (AIM) with prominent B cell aggregates (BCM) on muscle biopsy is an uncommon finding. We aimed to compare the characteristics and clinical course of patients with BCM on muscle biopsy to those without and characterize B cell infiltrates in the muscle of these patients.

We performed a retrospective study of subjects with AIM in the Canada Inflammatory Myopathy Study (CIMS) cohort to identify cases with BCM on muscle biopsy, which was defined as ≥30 CD20+ cells/aggregate. AIM cases without BCM that encompassed the broader spectrum of AIM, namely dermatomyositis, overlap myositis and inclusion body myositis were selected as controls. Descriptive statistics were used to compare BCM cases and controls. Cyclic immunofluorescence (Cyc-IF) was performed to characterize inflammatory infiltrates and B cell structures. We included 69 subjects (mean age at diagnosis 51±16 years, 77% females): 22 BCM, 24 dermatomyositis, 14 overlap myositis, and inclusion body myositis. Most BCM subjects (18/22, 82%) had an associated autoimmune disease: 12 (55%) had systemic sclerosis, 5 (23%) rheumatoid arthritis and one (5%) systemic lupus erythematosus/systemic sclerosis overlap. Extra-muscular features found in BCM patients included arthritis (50%), interstitial lung disease (43%), Raynaud’s phenomenon (32%), and dermatomyositis rash (27%). Two patients (9%) had facial muscle weakness and one (5%) had positive anti-AChR autoantibodies. In BCM subjects, upper extremities were weaker than lower extremities in 7/21 (33%) of cases. Neck flexor weakness was frequent (17/22, 77%), while neck extensor weakness was uncommon (1/15, 7%). Cyclic immunofluorescence (Cyc-IF) spatial analysis of BCM biopsies displayed many features akin to tertiary lymphoid structures. Findings suggest possible involvement of both the traditional germinal center reaction and the extrafollicular pathway in BCM. In conclusion, in this series of myositis with B cell aggregates reported to date we found clinical similarities (i.e., associated with overlapping autoimmune diseases) and differences (i.e., muscle weakness distribution) with previous reports. The discovery of tertiary lymphoid structures on spatial analysis of muscle biopsies of BCM patients provides novel insight into its pathophysiology and potential therapeutic targets.

## 1. Introduction

### 1.1 Autoimmune inflammatory myopathies

Autoimmune inflammatory myopathies (AIM) are a group of heterogeneous diseases characterized by muscle inflammation and frequent extra-muscular features. Interaction between the immune system and skeletal muscle is central to the pathophysiology of AIM (1). Mononuclear immune cells, in particular T cells, are known to play an important role in the pathogenesis of AIM. CD4+ T-cells infiltrate the perivascular regions in dermatomyositis and are central to the development of the disease, while CD8+ T-cells, found predominantly in inclusion body myositis and polymyositis, cause muscle fiber destruction through MHC-1 recognition of their cognate antigen.

The role of B cells remains inadequately characterized in AIM. Indeed, while rare B cells have been described in AIM, B cell aggregates are an uncommon feature on muscle histopathology (2) (3). Such aggregates can range from B cell-rich clusters to organized, follicle-like structures (4) (5). In one of the earliest series described by Pestronk and al. (6), 10 patients with prominent perimysial and perivascular B cell aggregates were described. All patients had an associated autoimmune disease (i.e., rheumatoid arthritis, connective tissue disease and/or myasthenia gravis). Those patients had a predominantly proximal upper extremity and cervical pattern of weakness (brachio-cervical), with neck extensor involvement and relative sparing of neck flexors. Additional small case series have since been reported, sharing similarities with the Pestronk series (7) (8) (9) (10). Recently, an inclusion body myositis phenotype has been described in 5 of 8 patients with prominent B cells aggregates on muscle biopsy (11). Nonetheless, the literature still remains limited, consisting primarily of case series. There are significant gaps in recognizing the spectrum of their clinical presentation and the disease course of these patients and understanding the conditions allowing the muscle to be permissive for B cell infiltration in certain AIM patients.

### 1.2 Tertiary Lymphoid Structures (TLS)

In addition to the limited understanding of the significance of B cell infiltration in muscle tissue, the pathophysiologic role of B cell aggregates in muscle tissue is poorly described. B cells have been reported in other autoimmune diseases such as lupus nephritis and rheumatoid arthritis where they are known to participate in the germinal center reaction (12). This process leads to B cells increasing their immunoglobulin affinity through somatic hypermutation and isotype class switching to produce potent antibodies. Normally this process is restricted to lymph nodes in the context of infection, but they can appear in non-lymphoid tissue, in which they are called tertiary lymphoid structures. In this context, the tissue becomes permissive for many immune cells and acquires lymphoid organ functions. In immune-mediated diseases, this usually includes production of autoantibodies (12).

The objectives of this study were 1) to compare the characteristics of AIM patients with prominent B cell aggregates and controls and 2) to characterize the inflammatory infiltrates in muscle biopsies of AIM patients with prominent B cell aggregates and controls using cyclic immunofluorescence (Cyc-IF) (13).

## 2. Methods

### 2.1 Population

We conducted a retrospective case-control study of AIM patients enrolled in the Canadian Inflammatory Myopathy Study (CIMS), a multicenter prospective cohort of adult AIM patients recruiting in 6 centers across Canada. Participants included in this longitudinal registry are followed at yearly intervals and assessed with standardized evaluations (14), including detailed history and physical examination, self-reported questionnaires and laboratory investigations and the core set measures endorsed by the International Myositis Assessments and Clinical Studies (IMACS) group (14).

Subjects were included in this study if they underwent a muscle biopsy which was read by an experienced neuropathologist (JK, BE). All patients with B cell aggregates were included as cases. Controls without B cell aggregates were selected from the CIMS registry to encompass the clinical spectrum of AIM, including patients with dermatomyositis, overlap myositis and inclusion body myositis. Under current classification nomenclature, polymyositis has become a rare entity and thus was not been specifically included as a control group (15). Immune-mediated necrotizing myopathy patients were not included as control in this study as they typically have minimal or no lymphocytic infiltrates on muscle biopsy (16).

### 2.2 Classification

All patients were classified using the EULAR/ACR classification criteria for AIM (17). Patients were then grouped into BCM cases or controls. AIM with prominent B cell aggregates cases were defined as having ≥30 CD20+ B cells/aggregate on muscle biopsy, consistent with previous studies on myositis with B cell aggregates (11). Controls were defined as having <30 CD20+ B cells/aggregate on muscle biopsy.

In addition to subgrouping patients according to the presence of B cell aggregates, all patients were also subclassified into an AIM subset by consensus of an adjudication committee (MH, YT, OLC, VL) using clinico-sero-pathological phenotyping (18)(19)(20). The overlap myositis group included myositis overlapping with another connective tissue disease and anti-synthetase syndrome. The presence of an overlapping connective tissue disease was defined as the presence of myositis plus features of systemic lupus erythematosus, systemic sclerosis, Sjögren’s disease, mixed connective tissue disease or rheumatoid arthritis; either meeting classification criteria or strongly suggestive of that particular connective tissue disease (21)(22)(23)(24)(25)(26). In particular, a diagnosis of scleromyositis was made if an AIM patient exhibited features of systemic sclerosis, such as Raynaud’s phenomenon, telangiectasia, puffy fingers, sclerodactyly, digital pits/ulcers, abnormal nailfold capillaries on videocapillaroscopy and/or capillary basement membrane reduplication on histopathology, even if not fulfilling the systemic sclerosis classification criteria (27)(28). A diagnosis of anti-synthase syndrome was considered if the patient was positive for an anti-tRNA-synthetase autoantibody and displaying one or more of: myositis, Raynaud’s phenomenon, arthritis, interstitial lung disease (ILD), fever or mechanic’s hands (29). AIM patients were classified as dermatomyositis if they met the 239^th^ European Neuromuscular Center (ENMC) dermatomyositis classification criteria (30). A patient was diagnosed with inclusion body myositis in the presence of supporting clinical features (i.e., deep finger flexor and/or knee extensor weakness) and histological features in addition to meet at least one classification criteria for the disease (31).

### 2.3 Clinical data

Demographic data including sex, age and ethnicity were self-reported by study participants. Follow-up period was calculated from the symptom onset to the date of last follow-up or until withdrawal of consent. Muscle strength was assessed at baseline visit by using the Manual Muscle Testing of 8 muscle groups bilaterally (MMT8, score range 0-150, higher value corresponding to better strength). Cancer associated AIM was defined as cancer arising within 3 years of AIM diagnosis (32). The presence of extra-muscular features (i.e., rash, Raynaud’s phenomenon, sclerodactyly, arthritis, interstitial lung disease, myocarditis, sicca and gastrointestinal dysmotility) was reported as present or absent by the study physician. Nailfold capillary abnormalities were reported by an experienced videocapillaroscopist (GG) using standardized definitions (33)(34). Serum peak creatine kinase (CK) and C-reactive protein (CRP) levels were retrieved from electronic health records. Treatment(s) and response to treatment were recorded. Remission was defined as a global disease activity score of 0-1/10 on the myositis disease activity assessment tool (MDAAT) (35).

### 2.4 Serologies

A commercial line immunoassay was used to identify myositis and systemic sclerosis autoantibodies (Euroimmun, Lubeck, Germany). To improve the specificity of autoantibody results, immunoblots were correlated with anti-nuclear antibody (ANA) indirect immunofluorescence patterns on HEP-2 substrates (ImmunoConcepts Inc., Sacramento, CA, USA; Inova Diagnostics, San Diego, CA, USA), or by a second assay on a separate platform (e.g. ALBIA, immunoprecipitation) (36).

### 2.5 Histopathological evaluation

Muscle biopsies were analysed at neuropathology referral centers by expert neuropathologists (JK, BE). Histopathological analyses were based on the ENMC recommendations for histological assessment of inflammatory muscle tissue (37), including hemotoxylin-eosin (H&E), hematoxylin-eosin-saffron (HES), Gomori trichrome, Congo red, NADH-TR, COX-SDH, acid phosphatase, alkaline phosphatase and non-specific esterase, all performed on 10 µm-thick fresh frozen sections. Immunohistochemical stains were performed on either a Ventana (JK) or DAKO (ES) platforms. Frozen or formalin-fixed paraffin-embedded sections were stained for membrane attack complex (MAC), MHC class I (MHC I), CD4 (helper T-cell), CD8 (cytotoxic T-cell), CD20 (B cells) and TDP43. When additional cell types were suspected on H&E, CD68 (macrophages) and CD138 (plasma cells) staining was performed. In select cases, electron microscopy was used to assess for capillary tubuloreticular inclusions and prominent capillary basement membrane reduplication (27)(38).

Lymphocytic infiltrates (endomysial, perimysial and perivascular), necrosis, MAC (sarcolemmal and capillary) and MHC I deposition were scored from 0 to 3 (0: absent, 1: focal, 2: moderate, 3: strong). Prominent B cell aggregates were further classified into “B cell rich” (30-99 CD20+ cells/aggregate), or “follicle-like” (≥100 CD20+ cells/aggregate), as reported by the neuropathologist. The localisation of such aggregates was also recorded.

### 2.6 Cytokine, chemokine and growth factor panel

A commercial Millipore human cytokine/chemokine 96-plex discovery assay® was performed using the Luminex® 200™ platform (Eve Technologies, Calgary, AB, Canada) at baseline research visit (or year 1 if unavailable) at a visit closest to their baseline visit in the CMS registry in BCM participants and select controls. In addition, thrombospondin 1 was tested in all samples (9). Given that there is no established reference interval for protein values, cytokines/chemokines were compared relatively to other samples in the same batch. Furthermore, results were analysed as: 1) clusters of co-expressed cytokines, and 2) cytokines involved in B cell maturation, proliferation and survival (39). Patients could be either treatment-naïve or on immunosuppressive treatment at the time the sera were collected. Samples with significant test interference were excluded from the study.

### 2.7 Cyclic immunofluorescence

Cyclic immunofluorescence was performed on available muscle biopsies, including 17 AIM with B cell aggregates, 11 DM patients, 10 OM and 7 IBM patients. Muscle biopsies were fixed in formalin, embedded in paraffin and cut into 4um thick sections by microtome at the University of Sherbrooke’s histology and electronic microscopy platform. Muscle biopsies were processed and stained with a comprehensive panel of 22 markers to identify subsets of infiltrating immune cells. A list identifying all the commercially conjugated and unconjugated antibodies used in this experiment is provided in **Supplementary Table 1,** with their respective clone, company of origin and dilution. For unconjugated antibodies, visualisation of the staining was performed with secondary labelling using Alexa Fluor conjugation kits according to the manufacturer’s protocols: Alexa Fluor 488 (Cat. A37570), Alexa Fluor 555 (Cat. A37571), Alexa Fluor 647 (Cat. A37573), and Alexa Fluor 750 (Cat. A37575).

Cyc-IF was performed according as previously described (13) (40) (41). In brief, muscle biopsy slides were deparaffinized using xylene. Antigen retrieval was then performed in pH6 citrate buffer inside a pressure cooker (Cuisinart CPC-600), for a duration of 20 minutes (high setting). After cooling, slides were rinsed in distilled water before being incubated in pH 9 Tris/EDTA buffer for a further 15 minutes and then blocked in PBS (10% normal goat serum and 1% BSA). Microscopy was performed using a ZEISS Axioscan 7 fluorescence slide scanner. First, autofluorescence was acquired before sequentially staining 7 rounds of markers. Each round comprised staining with a set of 4 antibodies conjugated to Alexa-Fluor 488, 555, 647 or 750 or similar fluorescence conjugates (with overnight incubation), an acquisition step and a quenching step using 3% hydrogen peroxide and 20 mM NaOH in PBS for 30 min. Signal quenching was verified before proceeding to the next round in order to ensure the quality of the signal observed.

Image processing integrated both established tools and custom python scripts. Image registration for the multiple rounds was done using ASHLAR (Alignment by Simultaneous Harmonization of Layer/Adjacency Registration). The complete methodology is available on GitHub (https://github.com/labsyspharm/ashlar). Segmentation and feature extraction (nucleus x and y coordinates, nucleus features, marker intensity) were then performed using QiTissue Image analysis software version 1.4.0 (Quantitative Imaging Systems, LLC, Portland, OR, USA).

To enable accurate tissue analysis and spatial interaction mapping, germinal centers were first identified and excluded from the dataset, as their densely packed cellular architecture can distort signal quantification from the rest of the follicle and compromise downstream analyses. Moreover, germinal centers are terminally differentiated structures and their special characteristics at least for the dark zones and light zones are well known and would not contribute further to our comprehension of the immune dynamic at play in the muscle biopsies.

Germinal center Dark zones were thus excluded during the segmentation based on Ki67, CD20 and CD3 expression. Their composition was thus analysed separately.

Extracted features were average cell intensity for each marker and individual cell coordinates. Cells were filtered based on size and autofluorescence levels, and autofluorescence was subtracted for each individual cell. Finaly marker expression normalization was performed using z score calculations across all samples to ensure that the signal could be compared from sample to sample (github: https://github.com/biodev/cycIF-workflow/tree/v1.0).

Marker expression was then used to phenotype the cells for subsequent analyses. Cells were initially classified into 5 major types: muscle (myocytes), B lymphocyte, T lymphocyte, macrophages and dendritic cells. Markers used for this step included desmin for myocytes, CD19, CD20, CD11c, CD27, CD138, IGD, PD1 and CXCR5 for B cells, CD3, CD4, CD8, CD45RO, CD45RA, PD1, HLADR, CXCL13 and CXCR5 for T cells, CD68, CD163, CD169 for macrophages and CD11c, CD86 and HLA DR for dendritic cells. Exclusive markers were classified first to avoid mislabelling cells, and shared markers were used last in conjunction with lower thresholds of exclusive markers. Cells where then reclassified into subtypes using these markers.

Spatial analysis was performed based on the spatial coordinates from the segmentation data (‘Nuc_X’ and ‘Nuc_Y’). This data was converted from pixel to micrometers using a conversion factor (1 pixel = 0.346 micrometers). These coordinates where used to analyze the spatial distribution of different immune cell types around B cells using Euclidean distance measurements. Cyc-IF data analysis was performed using python version 3.10.15 implemented on Visual Studio Code version 1.101.2 via anaconda distribution. The pipeline calls for several specialized python libraries: Numpy version 1.23.5 and Pandas version 2.2.2 for data manipulation Matplotlib version 3.9.2 and Seaborn 0.13.2 for generating figures. Spatial analysis was performed based on Euclidean distances, using spatial analysis libraries such as Sklearn version 1.5.1 and Shapely version 2.0.5.

Validation of the inflammatory nature of the muscle biopsy samples was achieved by identification of both macrophage and T lymphocyte clusters known to be found in AIM. Macrophages and T lymphocyte infiltrates have been reported across all auto-immune myopathies as a hallmark pathological feature, used in the histopathological step in the diagnosis of these diseases (42,43). In the absence of clear quantitative definition of infiltration, clusters of 10 cells were used as validation threshold to check for presence of these infiltrates (44). Muscle biopsies where then reclassified based on the presence of B cell clusters: aggregates were defined as a cluster with more than 30 B cells and follicles were defined as clusters with more than 100 B cells, in accordance with the preliminary study from Meyer & al.(11). Clustering stability for these values was verified with adjusted rand index (ARI). This classification was then used to analyse the different B cell environments’ compositions and properties.

Enrichment analysis was performed by comparing cellular compositions of different structures between the BCM and non-BCM groups. To better understand how the cells present in the biopsies are positioned and interact with each other, neighborhood analysis was performed. Each B cell was identified as an origin to analyse the neighboring cells of each B cell, considering the structure to which the reference B cell belonged to. Neighborhood analysis around each individual B cell was achieved by analyzing which cells were enriched in the cell-to-cell interaction zone (0-75 μm) and which cells were enriched in the paracrine interaction zone (0-200 μm) for each type of structure. Cell interaction zones were extrapolated from multiple sources (45–48). If a cell subtype was enriched (BCM vs non-BCM) in both intervals, a second enrichment analysis was performed between the 2 zones within the myositis with B cell aggregates group to determine the area where the cell was most likely to exert its function.

### 2.8 Statistical analysis

Descriptive statistics were used for baseline characteristics (means, SD, medians, IQRs). Statistical analysis was performed using IBM SPSS version 27, python packages SciPy version 1.14.1 and StatsModels version 0.14.2. Comparisons between BCM cases and controls (non-BCM) were performed with the non-parametric Mann-Whitney U tests, whereas the Wilcoxon signed rank test was used for paired sample comparison within the groups. Standard statistical annotations where used: not significant (ns) for p ≥ 0.05, * for p < 0.05, ** for p < 0.01, and *** for p < 0.001. As statistical analysis were considered exploratory, and no correction was made for multiple testing.

### 2.9 Ethics statement

Ethics approval for this study was obtained at the Jewish General Hospital by the Research Ethic Board of the CIUSSS du Centre-Ouest-de-l’Île-de-Montréal in Montréal (#2023-3409). All subjects provided informed written consent to participate in the study.

## 3. Results

### 3.1 Baseline characteristics of AIM participants

We included 69 patients with AIM in the study, of which 22 had B cell aggregates. Amongst the AIM controls, 24 had dermatomyositis, 14 overlap myositis (8 scleromyositis, 4 anti-synthase syndrome, 1 mixed connective tissue disease and 1 undifferentiated connective tissue disease-overlap myositis) and 9 IBM (**Figure 1**). Baseline characteristics are presented in **Table 1**. The mean±SD age of diagnosis was 51±16 years, mean±SD duration of follow-up was 6±5 years and 53 (77%) patients were female. Most patients (53, 77%) were Caucasians, and 5 (8%) had cancer-associated AIM. The median (IQR) peak CK was 781 (247, 3553) U/L.

**Figure 1.**
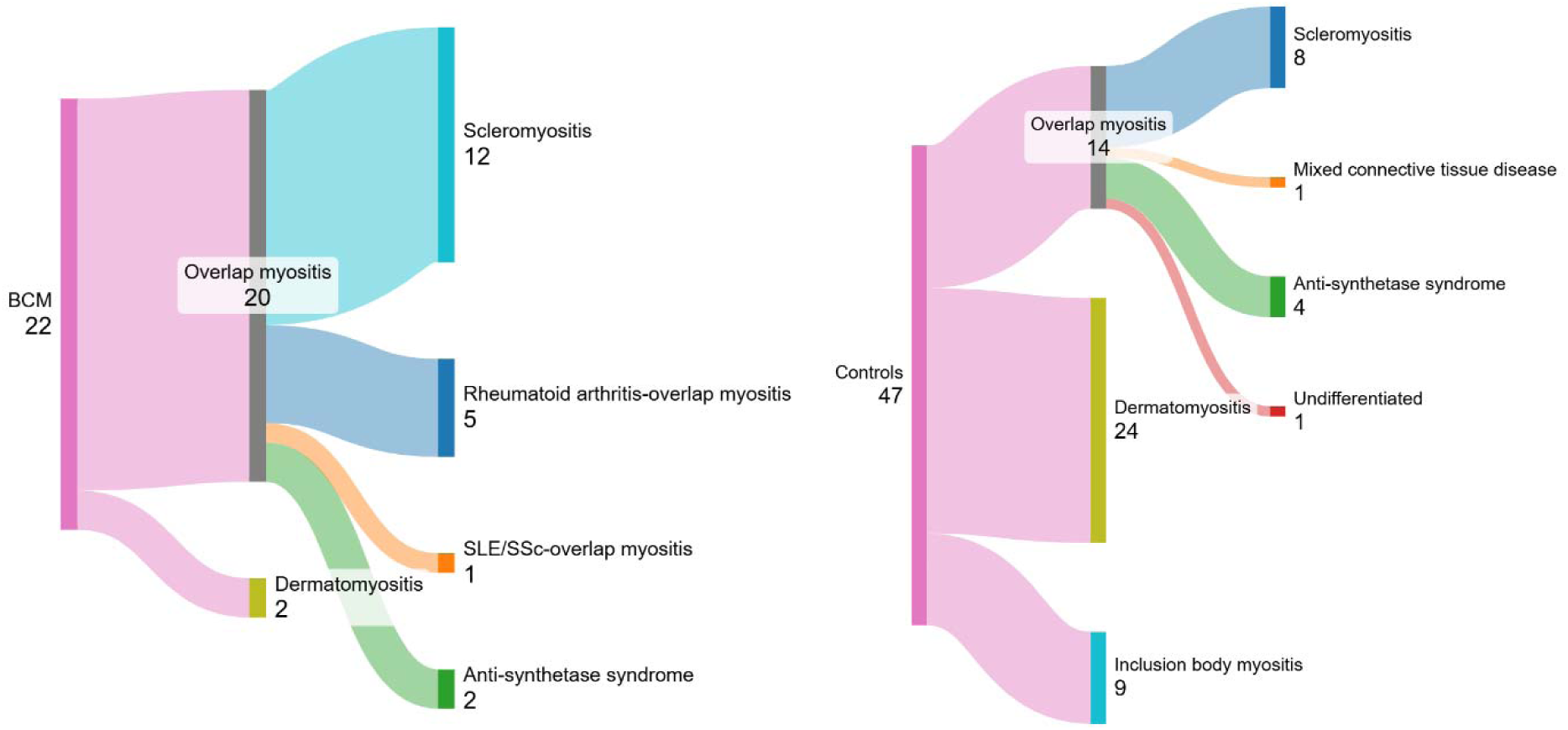
Distribution of 69 autoimmune myositis (AIM) patients into subgroups

**Table 1.**
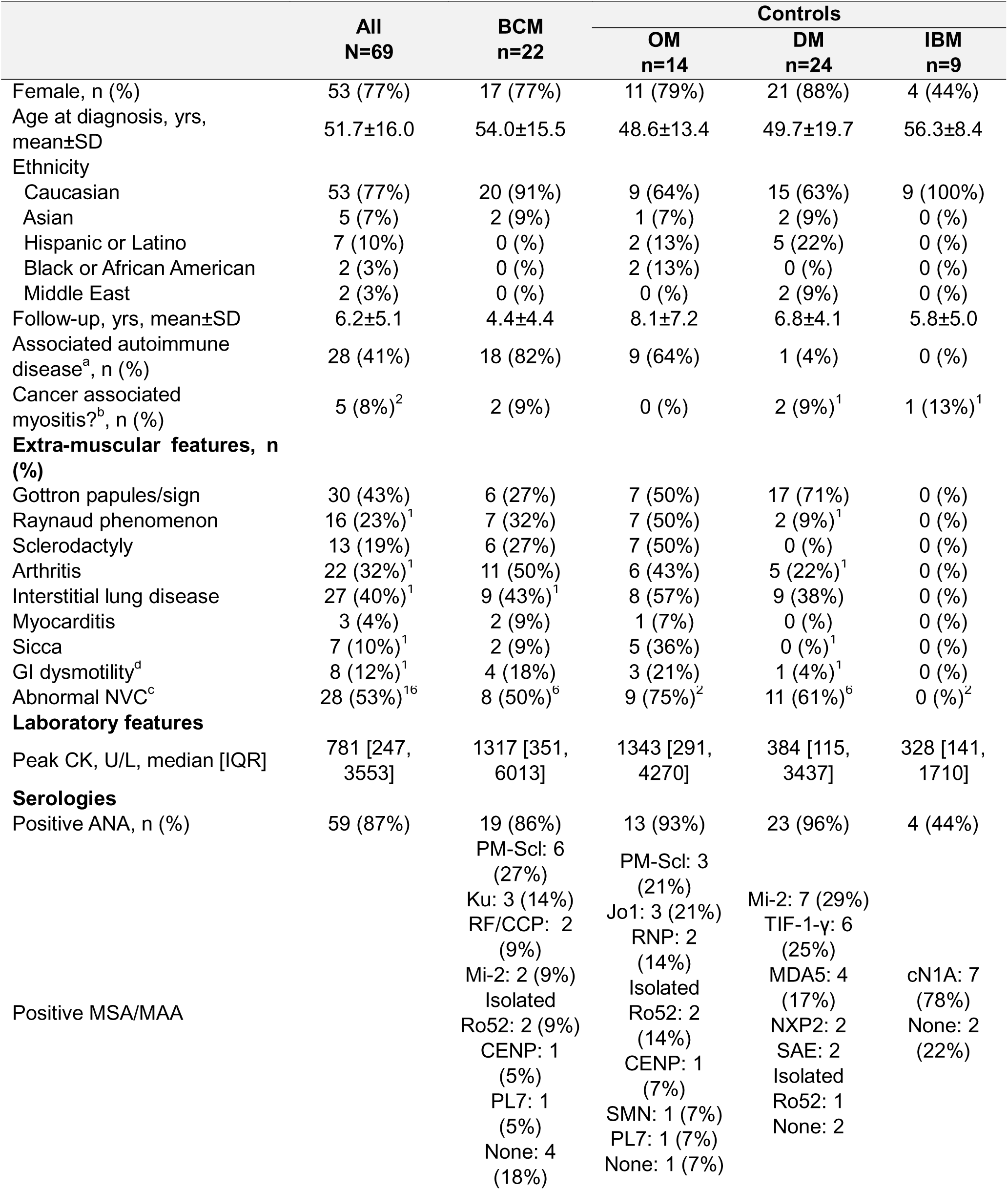
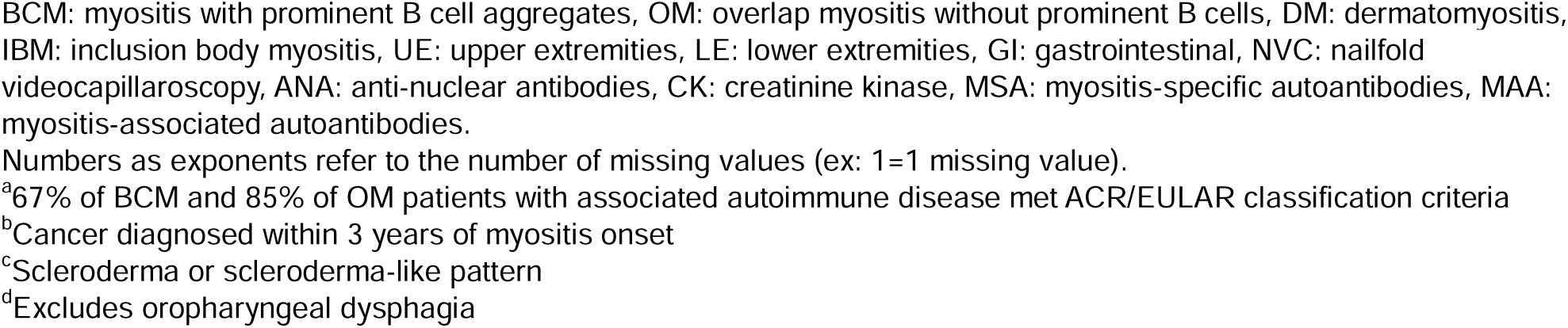
Baseline characteristics of 69 autoimmune myositis subjects.

Both the BCM and the overlap myositis groups had numerically more arthritis, Raynaud’s phenomenon, sclerodactyly and gastrointestinal dysmotility compared to the dermatomyositis and inclusion body myositis groups. Cytokine and chemokine profiles of 38 patients with available sera are presented in **Supplementary Figures 1 and 2**. No major differences in general and B cell stimulating cytokines were observed between the BCM cases and controls. Of note, 30 (80%) patients were on treatment, including 22 (58%) on prednisone, at the time the sera used for profiling.

**Figure 2.**
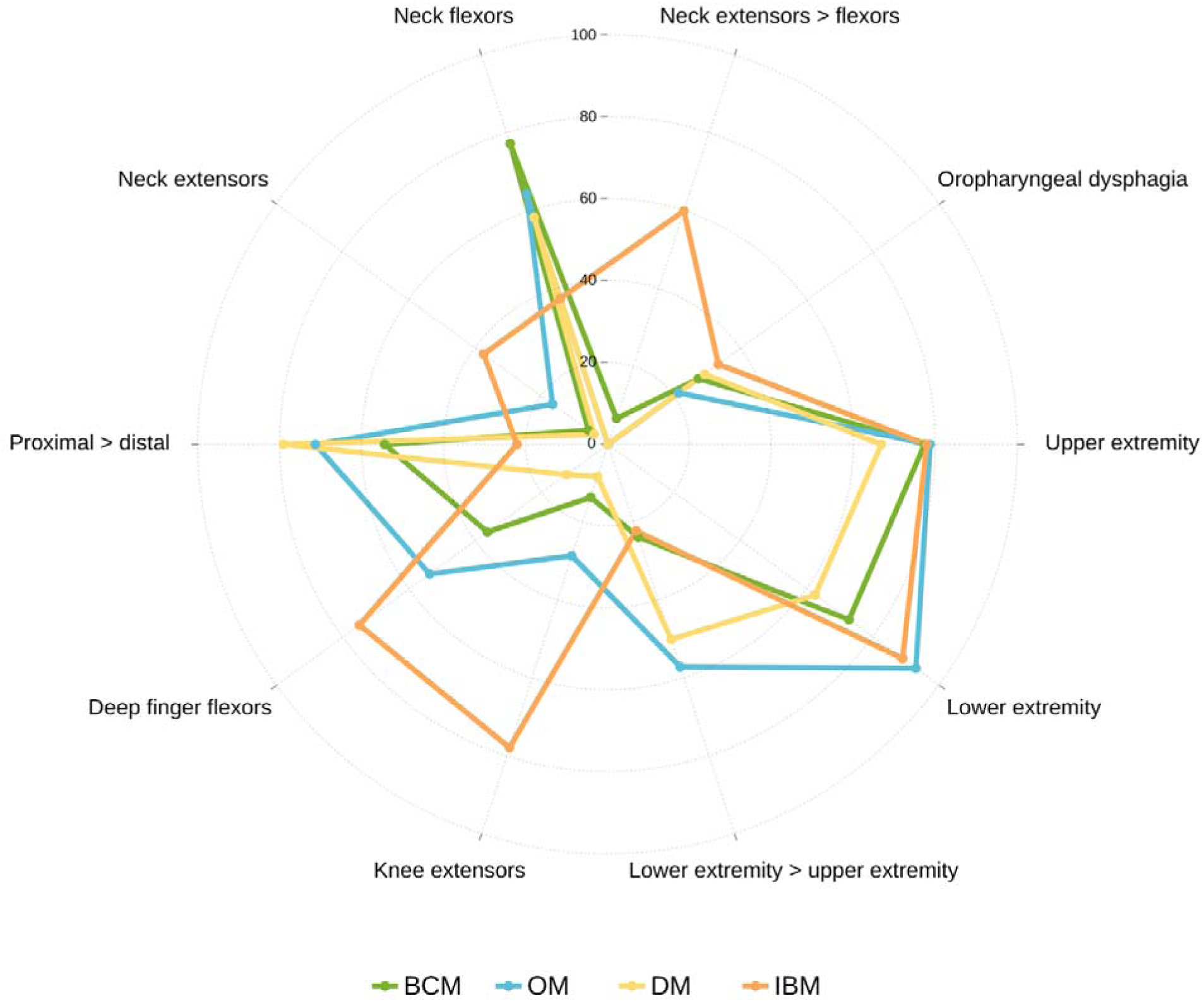
Pattern of muscle weakness across subgroups of autoimmune myositis BCM: myositis with prominent B cell aggregates, OM: overlap myositis without prominent B cell aggregates, DM: dermatomyositis, IBM: inclusion body myositis

### 3.2 Associated autoimmune diseases and autoantibodies

Most AIM patients with B cell aggregates on muscle biopsy (18/22, 82%) had an associated autoimmune disease: 12 (55%) had systemic sclerosis, 5 (23%) had rheumatoid arthritis and 1 (5%) had a systemic lupus erythematous/systemic sclerosis overlap. Eleven (61%) of these patients met ACR/EULAR classification criteria for those respective conditions (21)(24)(24,26). In the remaining 4 patients, 2 had anti-synthase syndrome and 2 had anti-Mi-2 dermatomyositis. Additional information is provided in **Supplementary Table 2.**

Eighteen (78%) patients with B cell aggregates in muscle tissue had positive myositis-specific or myositis-associated antibodies, namely anti-PM-Scl (6/22, 27%), -Ku (3/22, 14%), -RF/-CCP (2/22, 9%), -Mi-2 (2/22, 9%), isolated -Ro52 (2/22, 9%), -CENP (1/22, 5%) and -PL7 (1/22, 5%).

ANA was positive in 19 (86%), with speckled (12/19) and nucleolar (4/19) immunofluorescence patterns being the most common.

### 3.3 Pattern of muscle weakness

The pattern of muscle weakness of cases and controls is illustrated in **Figure 2**. Most patients with myositis with B cell aggregates had both upper (77%) and lower (73%) extremity weakness, with upper extremities weaker than lower extremities in a third of cases. We found no significant difference with the controls. Only one patient had weaker neck extensors compared to flexors. One quarter of the BCM cases were affected by oropharyngeal dysphagia. Two BCM patients had facial muscle weakness (one with positive AChR antibodies) and two had cranial nerve palsies.

Compared to the overlap myositis and dermatomyositis subgroups, BCM had less lower extremity dominant weakness (60%, 48% and 24%, respectively). While axial weakness was common in BCM 73%, only one patient (7%) had weaker neck extensors compared to flexors, compared to 60% in the IBM group. Compared to inclusion body myositis patients, BCM had less distal weakness (89% vs 50%), including deep finger flexors (75% vs 36%) and knee extensors (78% vs 14%). Two BCM patients met 2024 ENMC criteria for inclusion body myositis (31), although their overall presentation was not suggestive of this diagnosis.

### 3.4 Muscle histopathology

Muscle histology findings of 67 patients are detailed in **Supplementary Table 3**. Most biopsies were done in the deltoid (64%), with quadriceps (12%) and biceps (9%) being other common sites.

### 3.5 Management and treatment response of BCM patients

The management and treatment response of patients with B cells on muscle histology is presented in **Supplementary Figure 6**. Twenty (91%) BCM patients received glucocorticoids and eight (36%) patients received intra-veinous immunoglobulin (IVIG). The two patients who did not received glucocorticoids met classification criteria for systemic sclerosis.

The two most common first-line immunosuppressive treatment for BCM patients were methotrexate (10/22, 45%) and mycophenolate mofetil (6/22, 27%). Most BCM patients (13/21, 62%) responded to first-line treatment. Seven (32%) BCM patients received rituximab, a B cell depleting agent, but only one (5%) received rituximab as first-line therapy in combination with methotrexate. The proportion of patients who received rituximab over the course of their disease was greater in the BCM compared to the dermatomyositis and inclusion body myositis group (32%, 8% and 0%, respectively) but was similar between the BCM and overlap myositis groups (32% vs 29%). Four out of seven (57%) BCM patients who received rituximab had clinical improvement. Two of the three BCM patients who did not respond to rituximab also failed multiple other lines of therapy and had a clinically and pathologically advanced inclusion body myositis phenotype. The remaining patient had distal-predominant weakness with granuloma infiltration on muscle histopathology and noted subjective improvement with upadacitinib, a JAK inhibitor.

### 3.6 Spatial analysis

Cyc-IF was performed on 45 muscle biopsies, of which 2 were excluded because of tissue fixation failure between rounds. Validation of the inflammatory nature of the muscle biopsy samples was achieved by identification of both macrophage and T lymphocyte clusters known to be found in AIM (**Figure 3A**). Macrophage and T lymphocyte infiltrates have been reported across all AIM as a hallmark pathological feature, used in the histopathological step in the diagnosis of these diseases (42,43). In the absence of clear quantitative definition of infiltration, clusters of 10 cells were used as validation threshold to check for presence of these infiltrates (44). Results show that nearly all the patients presented with either type of cluster: 39 of the 43 patients (90.7%) presented with macrophage clusters, 36 of the 43 patients (83.7%) with T lymphocytes clusters and 39 out of 43 (90.7%) with both clusters, thus providing evidence that Cyc-IF is a valid method to analyse muscle biopsies.

**Figure 3:**
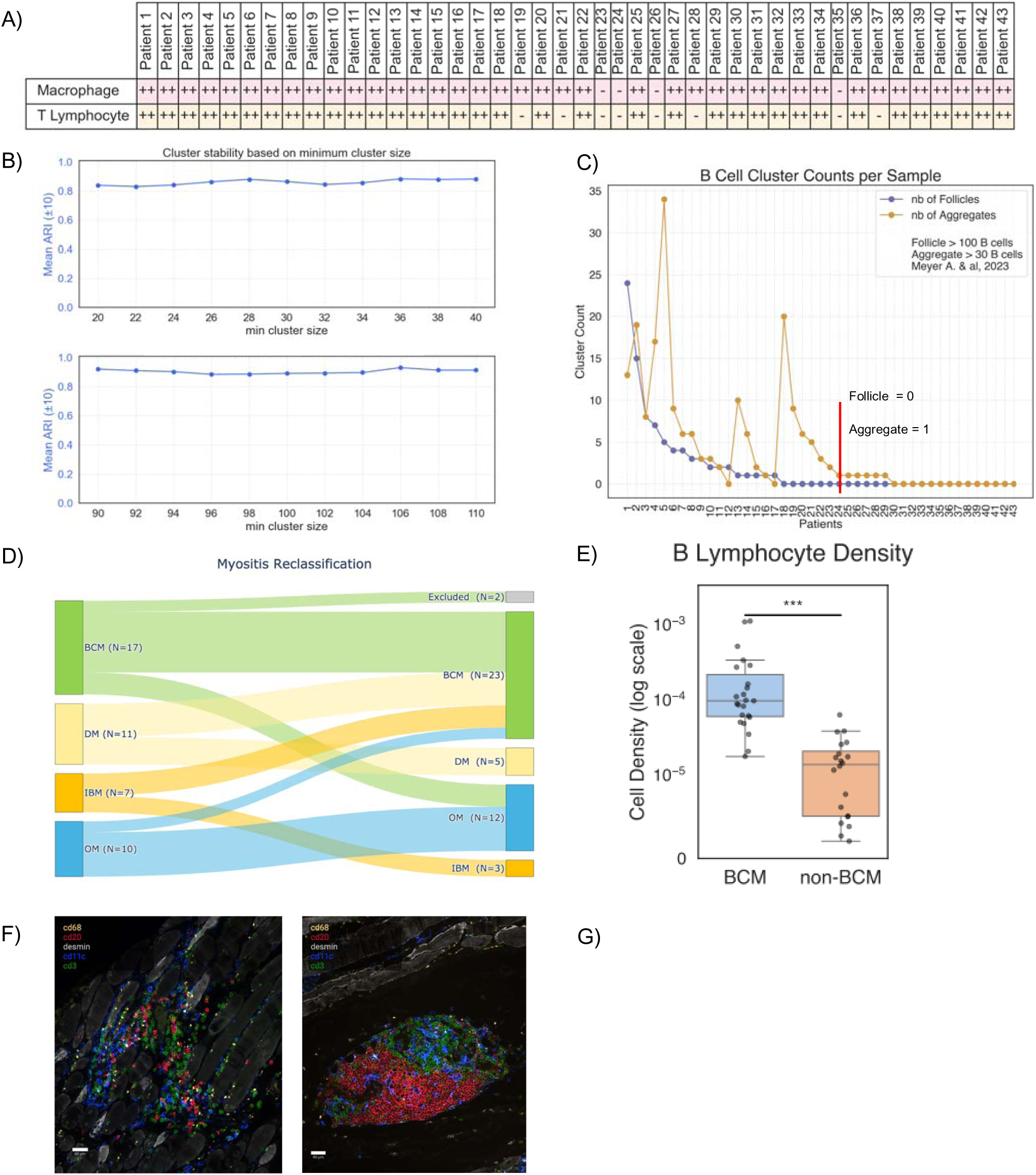
B cell based clustering **A)** Nearly all the patients presented with either macrophage or T cell clusters. **B)** clusters are stable for both the values (30 and 100 B cells) across a range of +/- 10 B cells, showing the initial definitions are valid (Adjusted Rand Index (ARI) > 0.8) **C)** BCM and non BCM groups where defined based on the presence of B cell clusters: BCM = >0 Follicles (>100B cells) or >1 aggregate (>30 B cells) **D)** patient reclassification based on Cyc-IF data **E)** BCM patients present higher overall B cell density than non-BCM patients) **F)** Image of a structure defined as aggregate (left) and a follicle (right).

Muscle biopsies where then reclassified based on the presence of B cell clusters: aggregates were defined as a cluster with more than 30 B cells and follicles were defined as clusters with more than 100 B cells, in accordance with the previous study from Meyer et al.(11).

The validity of this threshold was first mathematically validated using stability analysis. Validation was performed using Adjusted Rand Index (ARI) clustering stability measure on +/-10 B cells per cluster to determine the extent of the error coming from the clustering values. ARI is a clustering stability measure to determine the clustering sensitivity of a given parameter. We found an ARI above 0.8 showing that there was very little change between the clustering values of such a range, validating the usage of the definitions already published (49) (**Figure 3B**). Myositis with B cell aggregates was defined as having either more than 1 aggregate or the presence of a follicle. On the 43 analysable samples, 23 were then classified as BCM (53.5%) and 20 as non-BCM (46.6%) (**Figure 3 C, D**). In 7 cases, Cyc-IF detected the presence of TLS on muscle histology that was otherwise not observed using CD-20 staining.

The focus of this study was to understand which patients have the potential to organise B cell infiltration into organised functional structures like germinal centers as in other autoimmune diseases, compared to those presenting with non-specific B cell infiltration (50). This explains why a certain subset of non-BCM patients presented with higher B cell densities than some of their BCM counterparts even though overall BCM patients arbored significantly higher density of B cells in comparison to their non-BCM counterparts (**Figure 3 E**), confirming our current classification methodology.

Quantification of the infiltrates was achieved by comparing the densities and percentage of surface area of different type of structures (follicles: clusters of more than 100 B cells, aggregates: clusters of more than 30 B cells and muscle: the rest of the tissue surface) between the BCM samples and non-BCM samples in order to correct for tissue sample size and cluster size. This was achieved by comparing both density and percentage of area covered by different types of clusters. Densities were defined as absolute count of the structure type per unit of area, in this case number of aggregates over total sample area or number of follicles over total sample area. BCM patients presented a significantly higher density of both type of structures in comparison to their non-BCM counterparts. To correct for cluster size variability, percentage of total sample area covered by aggregates and follicles was compared between the 2 groups. Again, BCM patients presented a significantly higher percentage of total area covered by B cell clusters. This data supports the distinction of 2 AIM groups: 1 with prominent B cell clusters and the other without. For future reference, any tissue region which was neither a follicle, nor an aggregate, was considered muscle (**Figure 3 F**).

The different types of infiltrating clusters were then qualitatively analysed to reveal phenotypic differences between BCM and non-BCM patients. To do so, immune cell type densities (i.e., B lymphocytes, dendritic cells, macrophages, and T lymphocytes) for each type of structure (i.e., aggregate, follicle and muscle) were compared between the groups. BCM presented significantly higher densities of each of the immune cell types across all types of structures. Statistical analysis on follicles was not performed as they were completely absent in the non BCM group, and therefore no enrichment analysis was possible.

Consequently, subsets of each immune cell types were analysed to identify the specific subsets of cells enriched in the BCM group. This analysis revealed that double negative (DN) 1 B cells, DN 4 B cells, tissue resident macrophages and CD163 macrophages were enriched in aggregates whereas DN 2 B cells, DN 3 B cells, centroblasts, naïve B cells, plasmablasts, plasmocytes, switched memory (SM) B cells, unswitched memory (USM) B cells, activated DCs, myeloid DCs, tissue resident DCs, CD169 macrophages, Tph, activated CD4 T cells, activated CD8 T cells, memory CD4 T cells, naïve CD4 T cells and naïve CD8 T cells were all enriched in both muscle and aggregate regions of BCM patients when compared to their non-BCM counterparts (**Table 3, Supplemental Figure 3**). Follicle composition could not be compared since there were no follicles in the non-BCM group.

**Table 3:**
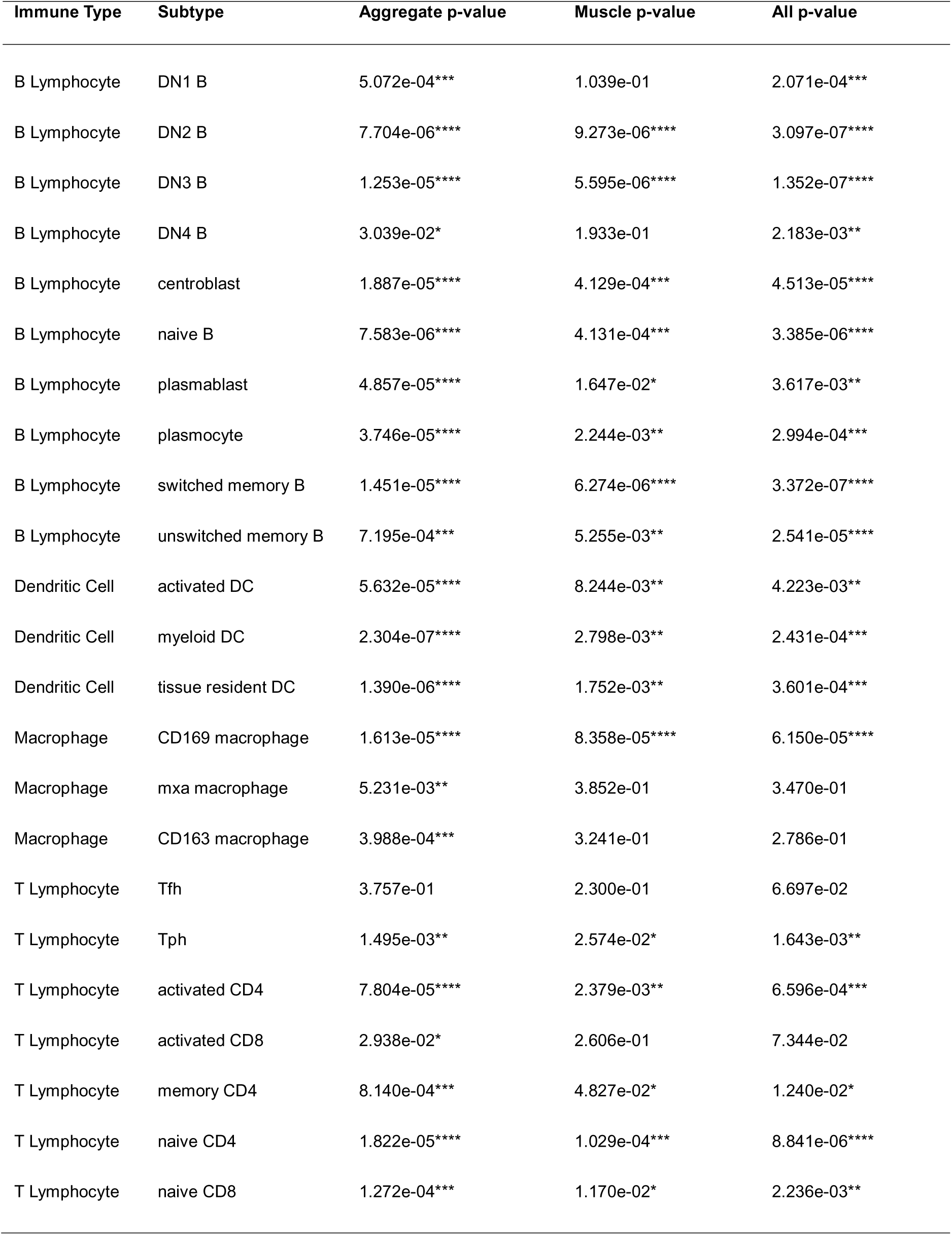
enriched cells in BCM group compared to non-BCM group, structure wise. All significantly enriched cells were enriched in the BCM group.

Neighborhood analysis around each individual B cell was achieved by analyzing which cells were enriched in the cell-to-cell interaction zone (0-75 μm) and which cells were enriched in the paracrine interaction zone (0-200 μm) for each type of structure. Cell interaction zones were extrapolated from multiple sources (45–48). If a cell subtype was enriched (BCM vs non-BCM) in both the intervals, a second enrichment analysis was performed between the 2 zones within the BCM group to determine the area where the cell was most likely to exert its function. **Table 4 and Supplementary Figure 4** show the results from both analyses (BCM vs non-BCM and subsequent intra-BCM zone comparisons). These show an enrichment of DN1, DN2, DN3, DN4, USM B cells, SM B cells, memory CD4 T cells, naïve CD4 T cells, tissue resident DCs and activated DCs in the 0-75 μm cell to cell interaction zone and Tph, activated CD8 T cells, activated CD4 T cells, naïve B cells, plasmablasts and plasmocytes in the 0-200 μm paracrine interaction zone in the regions defined as muscle of the BCM patients compared to those regions of the non-BCM patients. In the aggregates, DN1, DN2, DN3, DN4, USM B cells, SM B cells, centroblasts, plasmablasts, plasmocytes, memory CD4 T cells, activated CD4 T cells, naïve CD4 T cells, activated CD8 T cells, naïve CD8 T cells, tissue resident DCs, activated DCs and CD169 macrophages were enriched in the 0-75 μm interaction zone and Tph and MxA macrophages were enriched in the 0-200 μm interaction zone.

**Table 4:** Distance to B cell interval based enrichment of immune subsets, NE = not enriched In the proximity of B cells, the cell types are enriched in the BCM phenotype.

| Subtype | Aggregate enrichment bin | Follicle enrichment bin | Muscle enrichment bin |
| --- | --- | --- | --- |
| DN1 B | 0-75 BCM | NE | 0-75 BCM |
| DN2 B | 0-75 BCM | 0-75 BCM | 0-75 BCM |
| DN3 B | 0-75 BCM | 0-75 BCM | 0-75 BCM |
| DN4 B | NE | NE | 0-75 BCM |
| centroblast | 0-75 BCM | NE | 0-75 BCM |
| naive B | 0-75 BCM | 0-75 BCM | 0-200 BCM |
| plasmablast | 0-75 BCM | 0-75 BCM | 0-200 BCM |
| plasmocyte | 0-75 BCM | 0-75 BCM | 0-200 BCM |
| switched memory B | 0-75 BCM | 0-75 BCM | 0-75 BCM |
| unswitched memory B | 0-75 BCM | 0-75 BCM | 0-75 BCM |
| activated DC | 0-75 BCM | 0-75 BCM | 0-75 BCM |
| tissue resident DC | 0-75 BCM | 0-75 BCM | 0-75 BCM |
| CD169 macrophage | 0-75 BCM | 0-75 BCM | 0-75 BCM |
| MxA macrophage | 0-200 BCM | 0-200 BCM | NE |
| CD163 macrophage | 0-75 BCM | 0-75 BCM | 0-75 non-BCM |
| Tfh | NE | NE | 0-75 BCM |
| Tph | 0-200 BCM | 0-75 BCM | 0-200 BCM |
| activated CD4 | 0-75 BCM | 0-75 BCM | 0-200 BCM |
| activated CD8 | 0-75 BCM | 0-200 BCM | 0-200 BCM |
| memory CD4 | 0-75 BCM | 0-200 BCM | 0-75 BCM |
| naive CD4 | 0-75 BCM | 0-75 BCM | 0-75 BCM |
| naive CD8 | 0-75 BCM | 0-75 BCM | 0-75 BCM |

Again, due to the absence of follicles in the non-BCM group, if a cell was found in the follicles of the BCM group, then the enrichment analysis was performed only within the BCM group, comparing the intervals. In the follicles, DN2, DN3, USM B cells, SM B cells, centroblasts, plasmablasts, Tph, activated CD4 T cells, naïve CD4 T cells, naïve CD8 T cells, tissue resident DCs, activated DCs, CD163 macrophages and CD169 macrophages were enriched in the 0-75 μm interaction zone and activated CD8 T cells, memory CD4 T cells, and MxA macrophages were enriched in the 0-200 μm interaction zone when comparing the 2 zones (**Table 4, Supplemental Figure 4)**.

Taken together, these results point to the fact that B cell aggregates in muscle biopsies of BCM patients are tertiary lymphoid structures.

## 4. Discussion

### 4.1 Autoimmunity and presentation of BCM patients

This study is the largest series of AIM with B cell aggregates reported to date and the first to compare BCM patients to AIM controls. In our cohort, we found different patterns of weakness as previously described, frequent association with autoimmune diseases and good treatment response to first line immunosuppression. We also found strong evidence suggesting the B cell aggregates in muscle biopsies are tertiary lymphoid structures, similar to those encountered in other autoimmune diseases.

In previous reports, a distinctive brachio-cephalic pattern of weakness was identified in patients with B cell aggregates on muscle biopsy (6–8). In our cohort, only 33% of patients had upper extremity predominant weakness. Similarly, while neck extensor more than neck flexor weakness was reported in the literature, this was not encountered in our study. Furthermore, only 2 of our patients had a distal, inclusion body myositis phenotype, whereas in the series reported by Meyer et al. (11), 5 of 8 patients met at least one classification criteria for inclusion body myositis. The differences between our results and those previously published could be due to referral bias (e.g., rheumatologist vs neurologists), time-lag bias and selection bias. Our findings suggest that the spectrum of AIM with B cell aggregates is broader than previously reported and that it can be difficult to distinguish a BCM and non-BCM subject based on clinical features alone.

Similar to other studies, most of our patients with B cell aggregates in muscle tissue had an associated autoimmune disease, with systemic sclerosis and rheumatoid arthritis being the most common associations in our cohort (6)(11). Similar B cell aggregates have been described in other chronic sites of inflammation, such as in the synovium in rheumatoid arthritis (51), in the lung in systemic sclerosis (52) and in salivary glands in Sjögren’s disease (53). TLS in tissue also seems to play a role in the pathogenesis of myasthenia gravis (19,54,55). However, it is important to highlight that B cell aggregates can also be found in other AIM subtypes, such as dermatomyositis and anti-synthase syndrome (5)(56)(57). Interestingly, in a subgroup analysis of the Rituximab in Myositis study, the presence of anti-Mi2 and anti-synthetase autoantibodies was predictive of a response to B cell depleting therapy(58) (59).

The presence of B cell aggregates in AIM presents a unique therapeutic opportunity to tailor care by identifying and targeting key cellular targets. In our study, while a third of BCM patients received B cell depleting therapy, it is nonetheless important to highlight that the majority of our BCM patients responded to first line immunosuppression (e.g., methotrexate), which might infer a good prognosis even without targeted therapy. On the other hand, two of our patients had an inclusion body myositis phenotype with highly suggestive muscle histology (i.e., endomysial inflammation, abundant COX-negative fibers, rimmed vacuoles, ragged-red fibers). Both of those patients were refractory to several lines of treatment and evolved similarly to sporadic inclusion body myositis patients. Management should thus be tailored on a case-by-case basis, and prospective studies are needed to determine the optimal management of BCM patients, including the early use of B cell targeted therapy. However, engaging in prospective studies for targeted therapy requires strong evidence of underlying pathomechanism.

### 4.2 Spatial analysis supports the presence of TLSs in BCM patients

B cells are central to the pathogenesis of autoimmune diseases, playing a role in autoantibody production, antigen presentation, upregulation of proinflammatory and profibrotic cytokines and interaction with other cells though cell-cell contact (60). While B cells are present across many subtypes of AIM, aggregates are uncommon. The localisation of B cells in muscle tissue is variable, with reports describing endomysial, perimysial and perivascular infiltration (6,61). B cells can also organise in a compartmentalized, follicle-like structure in the muscle in conjunction with a ring of T cells. They can thus behave as a tertiary lymphoid structure (TLS) and create a potential microenvironment for the perpetuation of immune dysregulation and autoimmunity (62). Multiparameter analysis using cyclic immunofluorescence supports this premise: first some clear tertiary lymphoid structure with typical structure were identified (dark and clear zone with concomitant ki-67 marking) and second most of the enriched cell subsets in BCM tissues were cells known to be involved in the germinal center reaction formation.

The cornerstone of TLS formation is the interaction between B and T cells: TLS rely on T cells for their establishment both at the level of the microenvironment and at the level of B cell stimulation to generate the B cell germinal center response. The former is achieved by specialized CD4 T cells called T peripheral helper cells (Tph) with migratory properties (CCR2 and CCR5) and are known to orchestrate the tertiary lymphoid structure response in autoimmune diseases and cancer (63). They secrete chemokines such as CXCL13, CCL19 and CCL21 which create the permissive microenvironment, thus enabling infiltration by various immune subsets namely large numbers of naïve lymphocytes (B and T) suggesting an acquired lymphoid organ function of the muscle (62,64). Presence of dendritic cells and CD169 macrophages suggest antigen presentation and activation of B cells (65,66) while colocalization of dendritic cells and naïve and activated CD4 T cells suggests in situ activation of T cells following antigen presentation (62,65,67–69). Memory CD4 T cells and activated CD4 T cells can then interact with B cells through CD40-CD40L to prevent excitotoxicity (B cell death in the absence of costimulation such as CD40-CD40L) and mitochondrial dysfunction following cognate interaction with an antigen or for isotype class switching (70,71). This is mirrored by the enrichment of centrobalsts (proliferating B cells that can undergo class switch and hypersomatic mutation(72)), which compose the germinal center dark zone (73). Enrichment of DN1 B cells (B cells which prematurely exit the germinal center reaction) suggests these TLSs are dysfunctional as double negative B cells are not found in healthy germinal centers. DN1 derived plasmablasts produce autoreactive antibodies due to their lack of specificity and perpetuate the inflammatory processes (74). In situ activation of B cells following muscle antigen stimulation might trigger formation of plasmablasts (short-lived) and plasmocytes (long-lived) which can undergo clonal expansion and might be responsible for the production autoreactive antibodies and perpetuation of the inflammatory processes. Switched memory B cells are also a product of germinal center reactions (74) and might contribute to clinical treatment response as they are known to migrate to the bone marrow, emphasizing the need for early detection and treatment of B cells that drive disease.

Other clues pointing at pathological germinal reaction include strong evidence of involvement of the extrafollicular pathway, as evidenced by DN2 and DN3 B cells, which are known to be pathogenic in SLE, IgG4-RD and COVID-19 infection (75–77). Their origin remains partially obscure, though DN2 B cells differentiate into plasmablasts following TLR7 stimulation (in SLE) and interact with Tph (74,78–80). DN3 cells are also thought to differentiate into short lived plasmablasts following commitment of B cells to the extrafollicular pathway (74). The remaining DN4 B cells are less studied but are thought to be IgE plasmocyte precursors (found in IgG4 disease) while their implication in other diseases still needs to be confirmed. In keeping with the clinical setting in which they were described, sustained expansion of DN B cells usually appear only in pathological situation (74). The importance of the extrafollicular pathway in BCM is also corroborated by interferon-related findings. In lupus nephritis, where TLS are frequently found, the proliferation of B cells is known to be mediated by type 1 interferon through promotion of B cell activating factor (BAFF) and A proliferation-inducing ligand (APRIL) (72) (81–83). In addition to previous reports of strong interferon signature in AIM, the heightened expression of MxA on macrophages, a surrogate for type 1β IFN activation, provides a strong case for a pathogenic role of interferon in BCM (84).

The first step for TLS formation is migration of immune cells in non-lymphoid tissue mediated by chemokines such as CXCL13. These chemokines are produced by Tph and have a paracrine effect on nearby immune cells: this is corroborated by the general location of Tph in the paracrine interaction zone in relation to B cells in BCM. As they are the epicenter of attraction, immune cells can understandably be very close to Tph in terminally differentiated structures even though the interaction is paracrine. This immune infiltration in the case of TLS is, as expected, disorganised without any clear B cell and T cell zone delimitation (85). The main mechanistic themes in these structures are antigen presentation as corroborated by the proximity of B cells to antigen presenting cells (65,66). Moreover, proximity of B cells to naïve CD4 T cells might suggest antigen presenting functions of B cells in TLS (74,86). Following activation, activated CD4 T cells and B cells need to interact to initiate a GC reaction which is corroborated by the proximity of these cells in B cells clusters (71). This will lead to differentiation of B cells into either the GC pathway (suggested by the presence of CD27+ class switched B cells) or the extrafollicular pathway, as observed in B cell clusters by the proximity to precursors and successors. As a matter of fact, enrichment of CD27-class switched B cells, typically resulting from a differentiation of B cells into extrafollicular pathway, suggests that the extrafollicular pathway of the germinal center has a role in the pathogenesis B cell myositis. This is something reported in other autoimmune diseases such as SLE (77,87). This corroborates the previously described interferon environment finding which is also present in SLE. The pathogenic role of interferon is also corroborated by the surprising infiltration of the aggregates and follicles by CD8 T cells. Usually, the follicle is devoid of CD8 T cells that are excluded from this anatomical region. Nonetheless, pathological germinal centers with such CD8 T cell infiltrates have been reported in some viral infections in the presence of high amounts of type 1 interferon (88,89). This observation might thus corroborate previous studies showing muscle interferon signature in AIM (90) (**Supplementary Figure 5**).

The absence of zone-specific enrichment of centroblasts in follicles can be explained by our deliberate exclusion of the dark zone for special analysis. As for the T follicular helper cells, the density of cells coupled with the antibody quality for CXCR5 can explain the lack of enrichment. This may be due to the phenotyping strategy which usually processes the specific markers first (marker signal specificity and strength coupled with cell type expression specificity). Since the samples are not single layers of cells and Tfh are usually found in highly dense areas, it is possible that those cells were mislabeled as the cell type of a cell underneath expressing a stronger and more specific marker. This is an inherent limitation of Cyc-IF or any immunohistochemistry technique using a hierarchical phenotyping strategy. Further study with additional markers such as BCL-6 could help to resolve this question.

In summary, this study provides compelling evidence that the B cell aggregates in muscle tissue represent TLS, akin to those observed in other autoimmune diseases. Through comprehensive immunophenotyping and spatial analysis, we demonstrated that the muscle tissue in BCM patients harbors all the cellular components necessary for TLS formation. These findings suggest that both germinal center and extrafollicular pathways are active in the muscle microenvironment, potentially contributing to local immune dysregulation, and opens new avenues for understanding the immunopathogenesis of AIM. Still, there are many questions left to be answered. The role of TLS in pathological antibody production, which can accumulate in myofibre and disrupt the function of key cellular target responsible for muscle homeostasis, is still unclear(91)(92). Additionally, the relationship between TLS and the interferon response, a key signaling pathway in lymphoneogenesis and in many subtypes of AIM (90), remains to be elucidated. Further staining panels related to interferon response as well as BCR repertoire analysis could enhance understanding of muscle TLS. Are they the source of antibodies and pro-inflammatory cytokines or simply a consequence of the inflammatory processes?

Limitations include the retrospective design of this study, in which case initial pattern of weakness and response to therapy may be subject to information bias. Furthermore, no patient received rituximab alone as first line therapy, therefore making it difficult to assess the direct efficacy of B cell depleting therapy. Interestingly, in 7 cases, Cyc-IF detected the presence of TLS on muscle histology that was otherwise not observed using CD-20 staining. This suggests that conventional pathology staining may underestimate the proportion of BCM patients, and that TLS may play a role in an even greater percentage of AIM patients than previously thought.

## 5. Conclusion

In the largest series of myositis with B cell aggregates to date, we found both similarities and differences compared to other series. Most patient had an associated autoimmune disorder, in particular systemic sclerosis or rheumatoid arthritis. In contrast to previous studies, the brachio-cervical pattern of weakness was not a dominant phenotype in our subjects, and the presentation was more heterogenous than previously described. In fact, in the absence of muscle histology, BCM patients can be clinically indistinguishable to non-BCM patients. We found strong evidence suggesting that B cell aggregates in muscle biopsies are TLS. However further studies are required to elucidate the significance of TLS on muscle biopsy, and the role of B cell depleting therapy.

## Data Availability

All data produced in the present study are available upon reasonable request to the authors

## AUTHOR CONTRIBUTIONS

MH, VL, HAC, ML, HSC and BR contributed to the conceptualization of the study.

VL, MH, OLC were involved in patient sample contribution.

BR, HSC, DJ, JK, VL, ML, HAC contributed to methodology

BR, HSC, ZG, CdCCO, MH, VL, ML, HAC contributed to data analysis

MH, VL, HAC, ML were involved in funding acquisition

HAC, VL, ML, MH were responsible for supervision.

BR and HCS wrote the original draft, all authors were involved in reviewing and editing the final figures and manuscript.

## Conflicts of interest / Divulgations

BR: none

HSC: none

MH: 3, 5, 9, 17

YT: 1, 3, 5, 6, 8, 9, 14, 15, 24, 25

OCL: none

GG: none

EOF: none

BE:

JK: none

CdCCO: none

DJ: none

ZG: none

ML: none

VL: 3, 10, 18

HAC: 1, 2, 3, 4, 6, 7, 9, 11, 12, 13, 16, 18, 19, 20, 21, 22, 23, 24, 26, 27

1. Abbvie
2. Amgen
3. Astra Zeneca
4. BMS
5. Boehringer Ingelheim
6. Celltrion
7. Daiichi Sankyo
8. Eli Lilly
9. Fresenius Kabi
10. Gilead
11. GSK
12. Hoffmann-La Roche
13. Janssen
14. Kezar Life Science
15. Kyowa Kirin
16. Mantra Pharma
17. Merck
18. Novartis
19. Neomed
20. Otsuka
21. Pfizer
22. Sandoz
23. Sanofi
24. Sobi
25. UCBs
26. Vielabio
27. Xencor

## Acknowledgement

We would like to highlight the contribution of Radhika Prabhune and Melanie Baniña to the CIMS database and the ethics approval process.

---

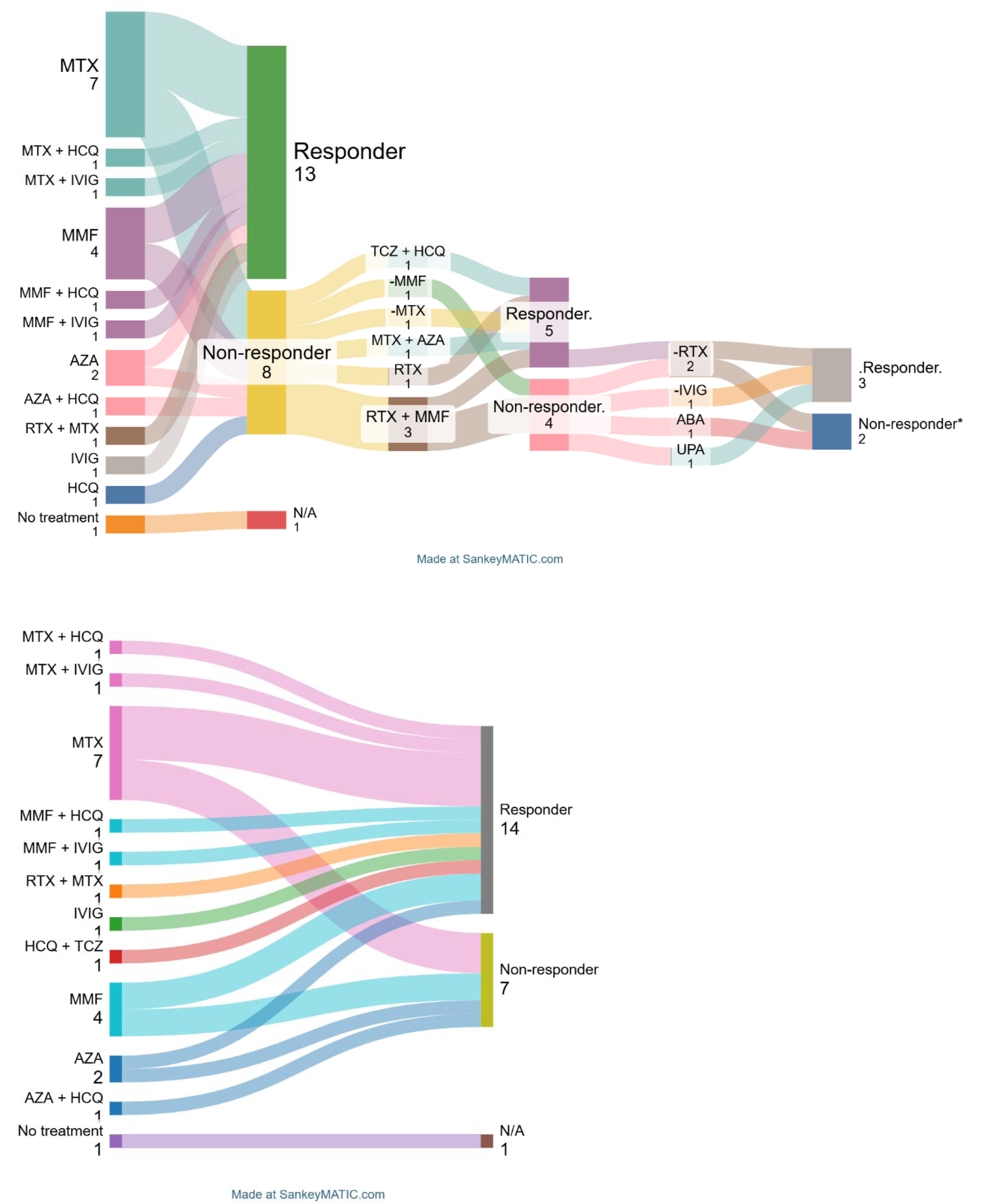

