## Supplemental figures for "Expanding the clinical spectrum of autoimmune inflammatory myopathies with prominent B cell aggregates: a case series"

**Supplementary Table 1**: Cyc-IF antibodies

| **Marker** | **Dilution** | **Company** | **Catalog number** | **Clone** |
| --- | --- | --- | --- | --- |
| CD3 | 1/50 | CST | 57869S | D7A6E |
| CD4 | 1/500 | ABCAM | ab196147 | EPR6855 |
| CD8 | 1/100 | LifeTechnologies | 50-0008-82 | AMC908 |
| CD11c | 1/50 | CST | 42756 | D3V1E |
| CD19 | 1/100 | ABCAM | ab274888 | EPR5906 |
| CD20 | 1/50 | ABCAM | ab198943 | EP459Y |
| CD27 | 1/100 | ABCAM | ab131254 | EPR8569 |
| CD45RA | 1/100 | BioRad | MCA88A488 | MCA88 |
| CD68 | 1/50 | Biolegend | 916104 | KP1 |
| CD86 | 1/25 | ABCAM | ab220188 | C86/1146 |
| CD138 | 1/50 | Novus | NBP2-89900AF647 | 160 |
| CD163 | 1/100 | ABCAM | ab281746 | EPR19518 |
| CD169 | 1/100 | ABCAM | ab306568 | SP216 |
| IgD | 1/25 | BioRad | STAR143F | polyclonal |
| CXCR5 | 1/25 | Biolegend | 356924 | J252D4 |
| CXCL13 | 1/50 | ABCAM | ab314944 | EPR23400-92 |
| PD1 | 1/50 | ABCAM | ab275126 | EPR4877(2) |
| Ki67 | 1/50 | Novus | NB110-90592 | polyclonal |
| Desmin | 1/500 | ABCAM | ab185033 | Y66 |
| HLADR | 1/25 | ABCAM | ab257320 | HLA-Pan/2967R |
| MxA | 1/100 | ABCAM | ab307342 | EPR24485-19 |

**Supplementary Table 2**. Clinical, serological and histopathological characteristics of 22 patients with myositis with prominent B-cells (BCM)

|  | **Classification** | | | | **Clinical features** | | | | | | | | **Muscle histopathology** | | | | | | | |
| --- | --- | --- | --- | --- | --- | --- | --- | --- | --- | --- | --- | --- | --- | --- | --- | --- | --- | --- | --- | --- |
| **Pt** | **Subclassification** | **Auto-antibodies** | **IIM EULAR/ACR criteria** | **Other CTD ACR/EULAR criteria** | **Upper vs lower extremity predominant weakness** | **Proximal vs distal predominant weakness** | **Axial weakness** | **Dysphagia** | **DM rash** | **Arthritis** | **RP** | **ILD** | **Inflammatory infiltrates** | **B-cell infiltrates, size and localisation** | **Necrosis** | **MHC1, localisation** | **MAC, localisation** | **Endothelial TRI** | **BM reduplication** | **Fibrosis** |
| **1** | OM (SM) | PM/Scl | Definite PM | Y | Same | Proximal | Y | N | N | N | Y | N | PV | PV | ++ | PF (+++) | Sarc (+), cap (+) | N | Y | N |
| **2** | OM (SM) | PM/Scl | Definite DM | Y | Lower | Proximal | Y | Y | Y | Y | N | Y | PV>EM, PMy | >100, PMy | ++ | PF (+++) | EM (+), cap (+) | N/A | N/A | N |
| **3** | OM (SM) | PM/Scl | Definite DM | Y | Lower | Proximal | Y | Y | Y | N | Y | Y | PV > EM, PMy | >30, EM | + | PF (+++) | Cap (++), PMy (+) | Y | N | N |
| **4** | OM (SM) | PM/Scl | Definite DM | Y | N/A | Proximal | Y | N | Y | N | Y | N | PV > EM, PMy | >100, EM | + | PF (+) | EM(++), cap (++) | Y | N | N |
| **5** | OM (SM) | PM/Scl | Not IIM | N | None | None | N | N | N | Y | N | Y | PV > EM, PMy | >30, EM | ++ | Cyto (+) | + | N/A | N/A | N |
| **6** | OM (SM) | PM/Scl | Definite PM | Y | Lower | Proximal | Y | N | N | Y | Y | Y | N/A | N/A | ++ | PF (+++) | Cap (+) | N/A | N/A | N |
| **7** | OM (SM) | CENP | Not IIM | N | Same | Same | Y | N | N | Y | N | N | PMy | >100, PV | N | PF (++) | Cap (++) | N/A | N/A | N |
| **8** | OM (SM) | Ku | Definite PM | Y | Upper | Distal | Y | Y | N | N | N | Y | EM > PMy, PV | Plasma cells +++ | ++ | EM (++) | Cap (++), sarc (+) | N | N | N |
| **9** | OM (SM) | Ku | Not IIM | N | None | None | N | N | N | Y | N | N | PMy, EM, PV | >100, PV | ++ | + | + | N/A | N/A | N |
| **10** | OM (SM) | Ro52 | Definite IBM | N | Upper | Distal | N | N | N | N | N | N | PMy, EM, PV | >100, EM | + | Sarc (++) | Cap (++), sarc (+) | N | N | N |
| **11** | OM (SM) | Seronegative | Definite IBM | Y | Upper | Distal | Y | Y | N | N | Y | N | EM, PV | >100, PV | ++ | Vasc (+++) | +++ | N | Y | Y |
| **12** | OM (SM) | Seronegative | Definite PM | N | Lower | Proxlmal | N | N | N | N | Y | Y | PM > PMy, PV | >30, EM | ++ | PF (+) | Sarc (+++), cap (++) | N/A | N/A | Y |
| **13** | OM (SLE/SSc) | Ku | Definite PM | Y | Upper | Proximal | N | N | N | Y | N | N | EM, PMy, PV | >100, EM | ++ | ++ | Sarc (++), cap (+) | N | N | N |
| **14** | OM (RA) | RF/CCp | Definite PM | Y | Upper | Proximal | Y | N | N | Y | N | N | EM > PV > PMy | >100. EM | +++ | EM (++) | EM (+, cap (+) | N/A | N/A | Y |
| **15** | OM (RA) | RF/CCP | Definite IBM | Y | Upper | Distal | Y | N | Y | Y | N | N | EM | >100, PV | + | Sarc (++) | Sarc (++), cap (+) | N/A | N/A | Y |
| **16** | OM (RA) | Seronegative | Definite PM | Y | Same | Proximal | Y | N | N | Y | N | N/A | EM > PV | >100, PV | + | EM (+++) | EM (+) | N/A | N/A | Y |
| **17** | OM (RA) | Seronegative | Definite PM | N | Upper | Proximal | Y | Y | N | Y | N | N | EM > PMy, PV | >100, EM | ++ | Focal (+) | Cap (++), sarc (+) | N/A | N/A | Y |
| **18** | OM (arthritis) | Seronegative | Probable PM | . | None | None | N | N | N | Y | N | N | PMy > EM, PV | >100, PMy | N | PF (+++) | Cap (++), sarc (+) | N/A | N/A | N |
| **19** | OM (ASyS) | PL-7 | Definite DM | N/A | None | None | Y | N | Y | N | Y | Y | EM | >100, EM | + | Cap (++) | Sarc (+++) | Y | Y | N |
| **20** | OM (ASyS) | Ro52 | Definite PM | N/A | None | Distal | Y | Y | N | N | N | Y | EM, PV | >100, EM | ++ | ++ | PF (+++), cap (+) | N/A | N/A | Y |
| **21** | DM | Mi2 | Definite DM | N/A | Lower | Proximal | Y | N | Y | N | N | N | PMy, PV > EM | >30, EM | +++ | PF (++) | Sarc (+++), cap (+++) | N/A | N/A | N |
| **22** | DM | Mi2 | Definite DM | N/A | Same | Proximal | Y | N | Y | N | Mp | Y | EM, PMy, PV | >30, PMy | + | PF (+++) | Sarc (++), cap (+) | N/A | N/A | N |

CTD: connective tissue disease, DM: dermatomyositis, RP: Raynaud’s phenomenon, ILD: interstitial lung disease, MHC: major histocompatibility complex, MAC: membrane attack complex, TRI: tubuloreticular inclusions, BM: basement membrane, SM: scleromyositis, PM: polymyositis, PV: perivascular, PF: perifascicular, sarc: sarcolemmal, cap: capillary, EM: endomysial, PMy: perimysial, OM: overlap myositis, SLE: systemic lupus erythematosus, SSc: systemic sclerosis, RA: rheumatoid arthritis, ASyS: anti-synthetase syndrome.

**Supplementary Table 3**. Muscle histopathological findings in myositis with prominent B cell aggregates (BCM) compared to controls

|  | **All**  **N=67** | **BCM**  **n=22** | **OM  n=14** | **DM**  **n=23** | **IBM**  **n=8** |
| --- | --- | --- | --- | --- | --- |
| **Muscle biopsy site, n (%)** |  |  |  |  |  |
| Deltoid | 43/64 (67%) | 12/20 (60%) | 12/14 (86%) | 16/22 (73%) | 3/8 (38%) |
| Biceps brachii | 5/64 (8%) | 3/20 (15%) | 0/14 (0%) | 1/22 (5%) | 1/8 (13%) |
| Quadriceps | 14/64 (22%) | 4/20 (20%) | 2/14 (14%) | 5/22 (23%) | 3/8 (38%) |
| Other(s) | 2/64 (3%) | 1/20 (5%) | 0/14 (0%) | 0/22 (0%) | 1/8 (0%) |
| **Inflammatory domain** |  |  |  |  |  |
| Endomysial lymphocytes (H&E) | 1.2±1.0 | 1.7±1.0 | 1.0±0.9 | 0.5±0.5 | 1.75±0.9 |
| Perimysial lymphocytes (H&E) | 0.6±0.8 | 1.1±0.9 | 0.2±0.4 | 0.5±0.5 | 0 |
| Perivascular lymphocytes (H&E) | 1.1±1.0 | 1.6±0.9 | 0.6±0.8 | 0.9±0.9 | 1.0±0.9 |
| Increased macrophages, n (%) | 34/66 (52%) | 17/22 (77%) | 6/14 (43%) | 10/23 (44%) | 1/7 (14%) |
| **Structural domain** |  |  |  |  |  |
| Perimysial pathology, n (%) | 26/66 (39%) | 10/22 (46%) | 2/14 (14%) | 14/23 (61%) | 0/7 (0% |
| Perifascicular atrophy, n (%) | 29/67 (43%) | 13/22 (45%) | 3/14 (21%) | 13/23 (57%) | 0/8 (0%) |
| Necrosis, score±SD | 1.21±0.9 | 1.6±0.8 | 1.0±1.0 | 1.1±0.9 | 0.8±0.7 |
| Type II fiber atrophy, n (%) | 27/65 (42%) | 13/22 (45%) | 6/14 (43%) | 8/22 (36%) | 3/7 (43%) |
| Rimmed vacuoles, n (%) | 9/67 (13%) | 2/22 (9%) | 2/14 (14%) | 0/23 (0%) | 5/8 (63%) |
| Vacuoles (non-rimmed), n (%) | 9/67 (13%) | 0/22 (0%) | 0/14 (0%) | 9/23 (39%) | 0/8 (0%) |
| IBM features^a^ |  |  |  |  |  |
| **Immunochemistry domain** |  |  |  |  |  |
| MAC, sarcolemmal deposition, score±SD | 1.1±1.0 | 1.6±0.9 | 0.7±0.7 | 0.8±1.0 | 0.7±0.8 |
| MAC, capillary deposition, score±SD | 1.2±0.9 | 1.4±0.7 | 0.8±0.6 | 1.3±1.0 | 0.6±0.5 |
| MHC-1, sarcolemmal, score±SD | 2.19±0.9 | 2.1±0.8 | 1.75±0.8 | 2.3±1.0 | 2.71±0.8 |
| BCM: myositis with prominent B-cell aggregates, OM: overlap myositis without prominent B cells, DM: dermatomyositis, IBM: inclusion body myositis, MAC: membrane attack complex, BM: basal membrane, TRI: tubuloreticular inclusions  ^a^: red-rimmed vacuoles, ragged red fibers, significant COX-negative fibers | | | | | |

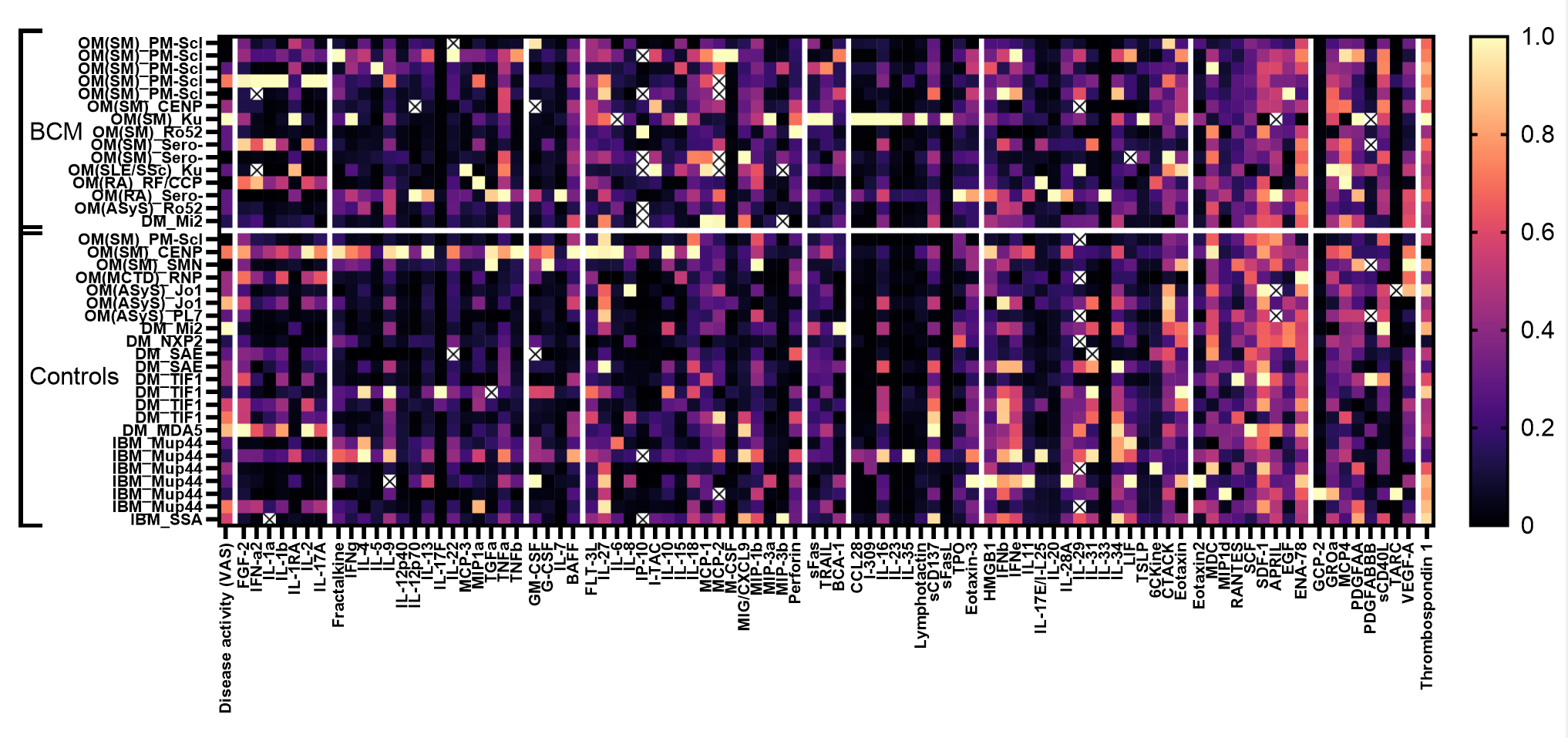
**Supplementary Figure 1**. Heatmap of cytokines and chemokines in 38* AIM patients.
BCM: myositis with prominent B-cell aggregates, SM: scleromyositis, OM: overlap myositis, DM: dermatomyositis, ASyS: anti-synthetase syndrome, IBM: inclusion body myositis, VAS: physician visual analog scale
Missing values are expressed as X.
*Two patients were excluded from the analysis because of outlier values.

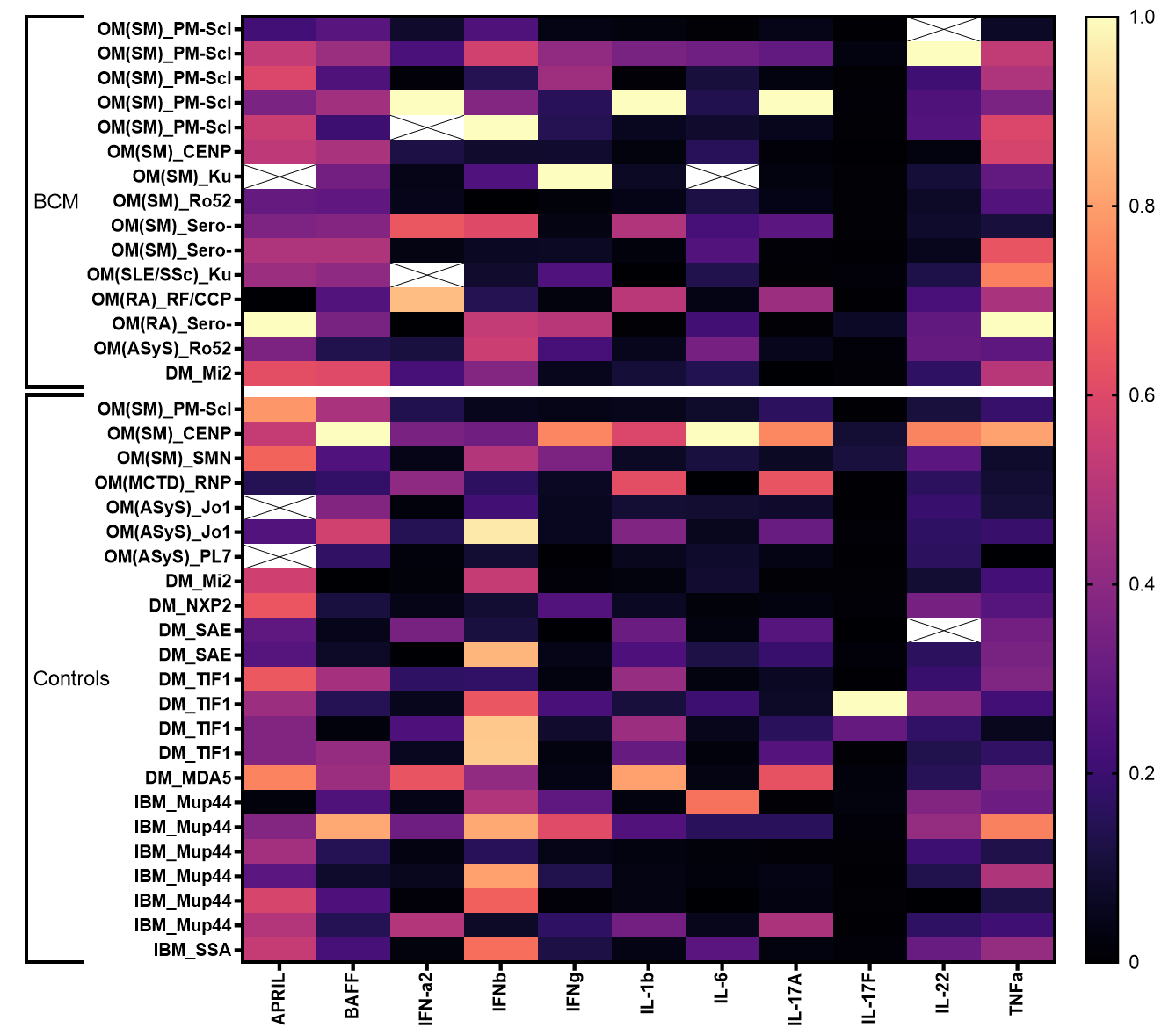

**Supplementary Figure 2**. Heatmap of cytokines involved in B-cell survival, proliferation and maturation in 38* AIM patients.
BCM: myositis with prominent B-cell aggregates, SM: scleromyositis, OM: overlap myositis, DM: dermatomyositis, ASyS: anti-synthetase syndrome, IBM: inclusion body myositis
Missing values are expressed as X.
*Two patients were excluded from the analysis because of outlier values.

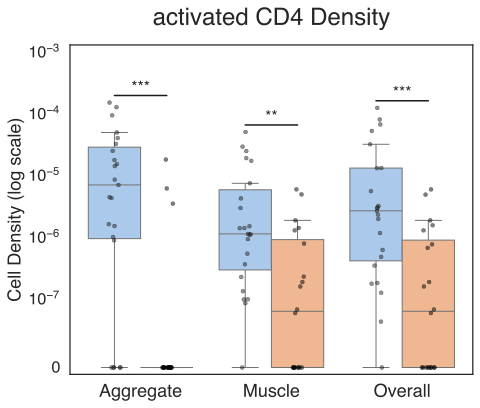

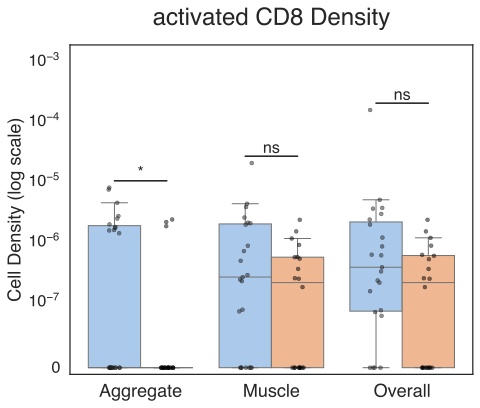

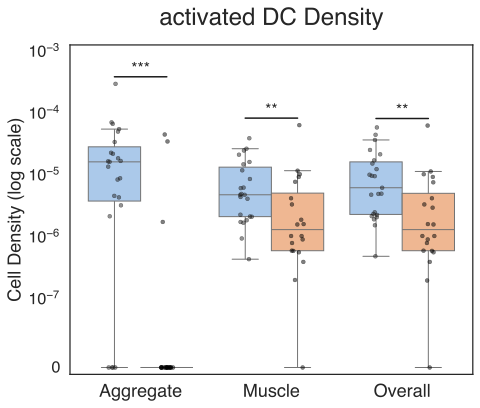

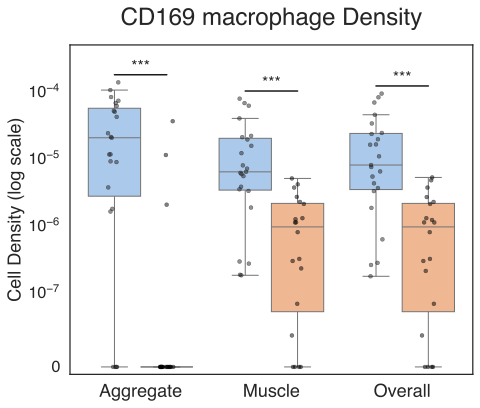

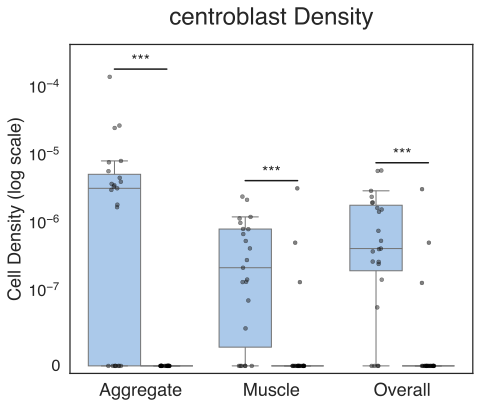

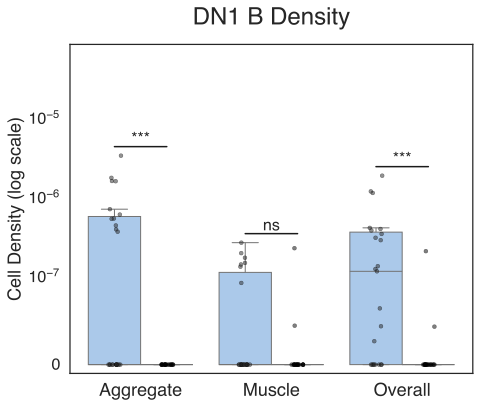

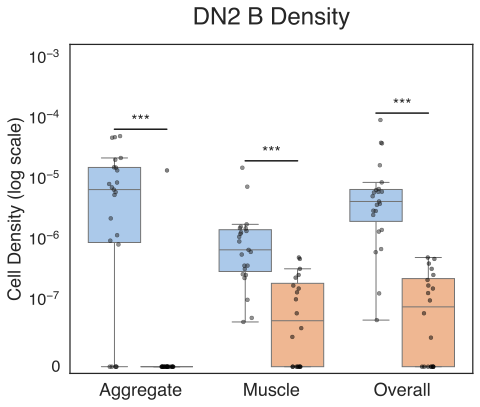

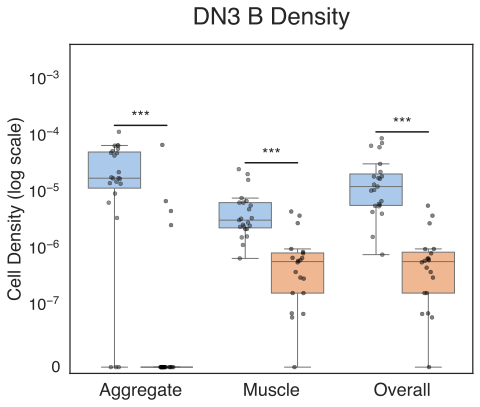

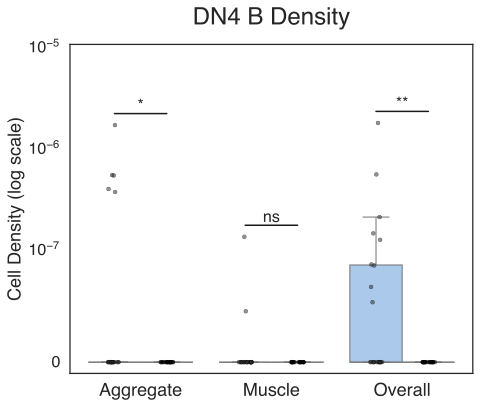

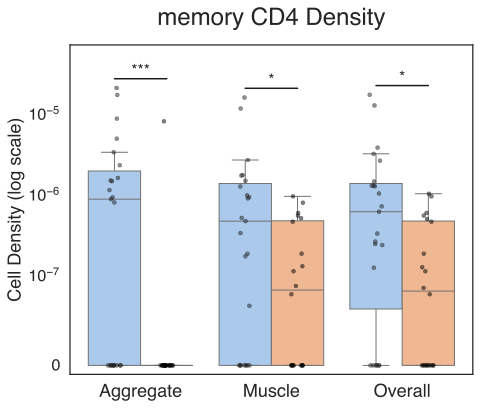

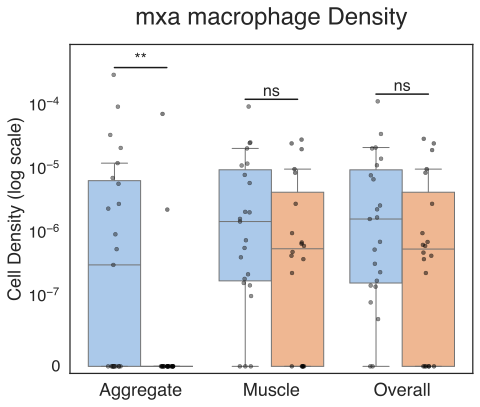

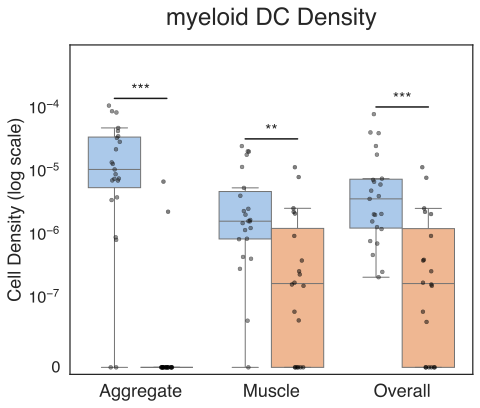

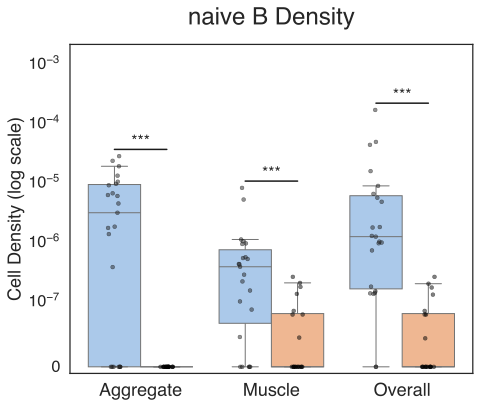

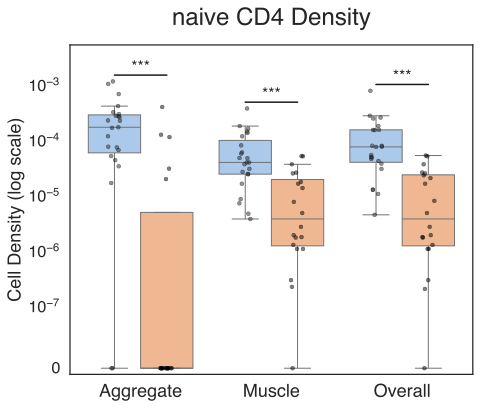

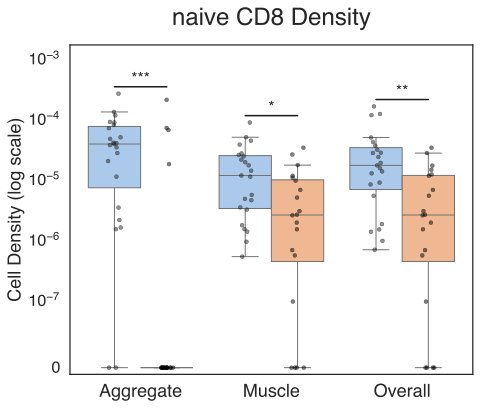

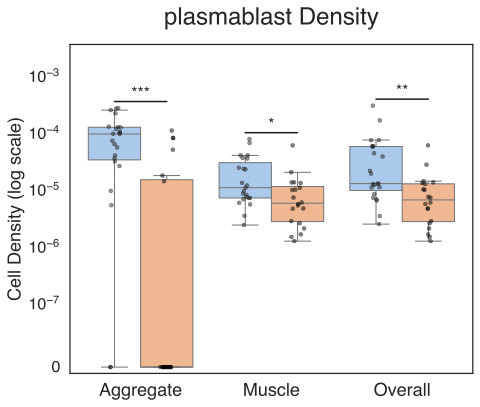

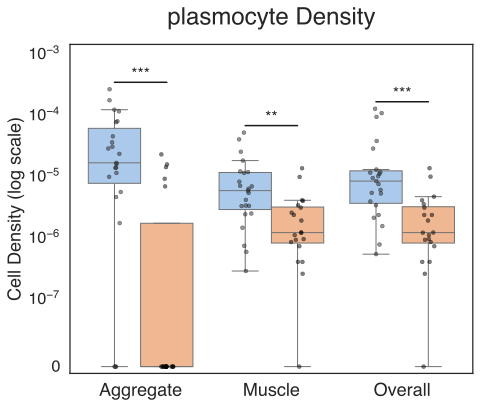

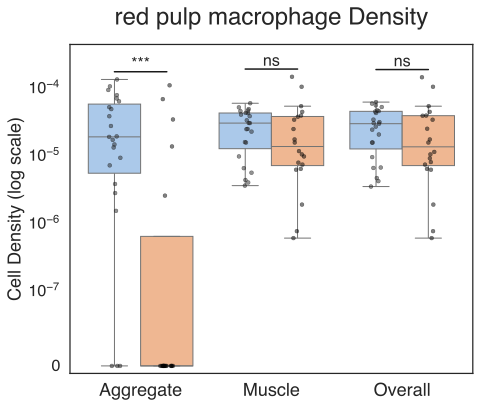

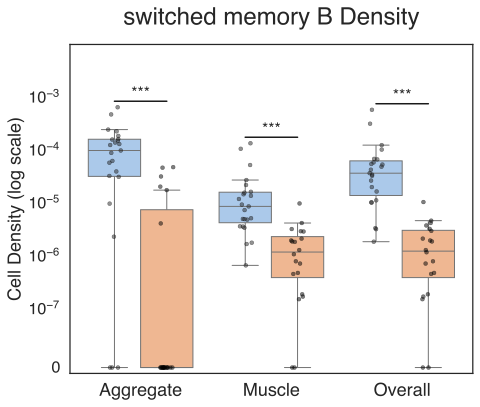

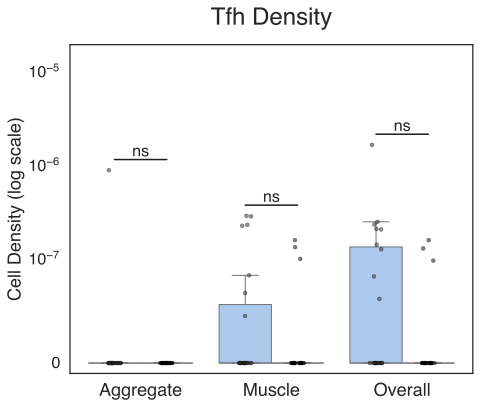

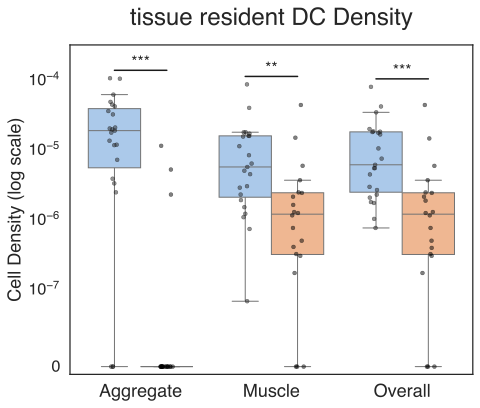

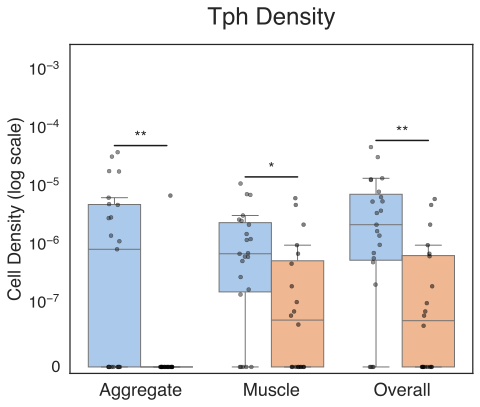

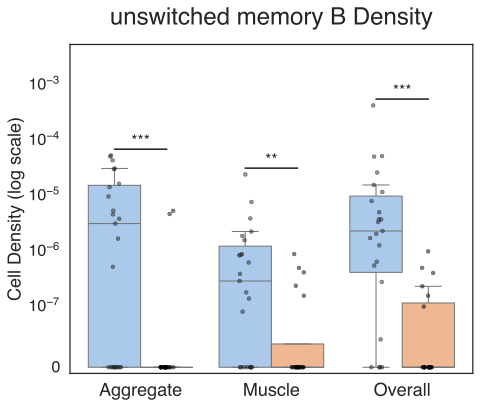

**Supplementary Figure 3 :** Enrichment of different immune cell types between BCM and non-BCM muscle tissue samples, for each type structure. (* : <0.05, ** : <0.01, *** : <0.001, ns: non-significant)

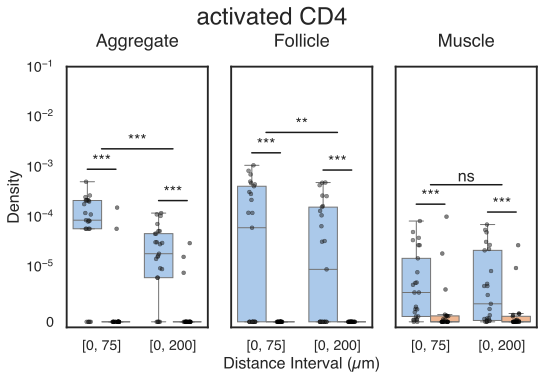

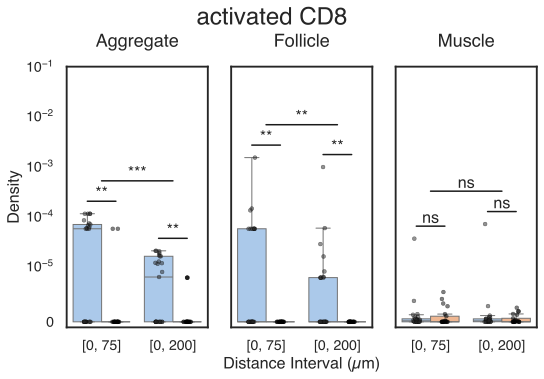

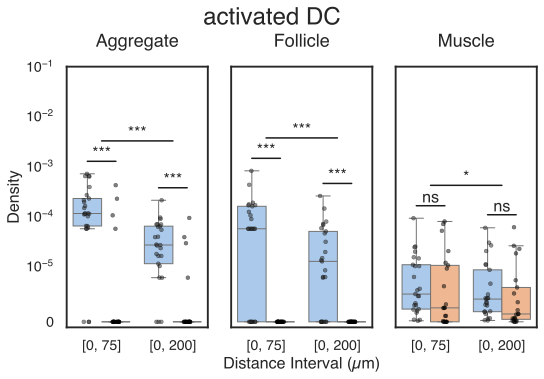

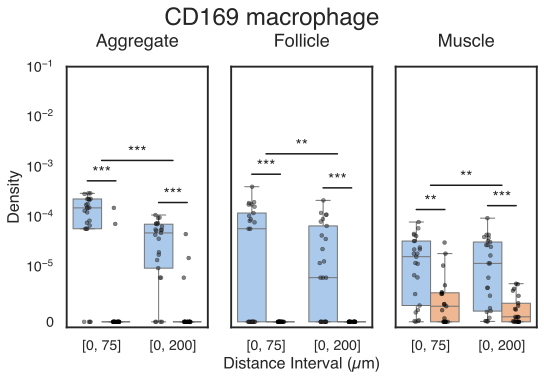

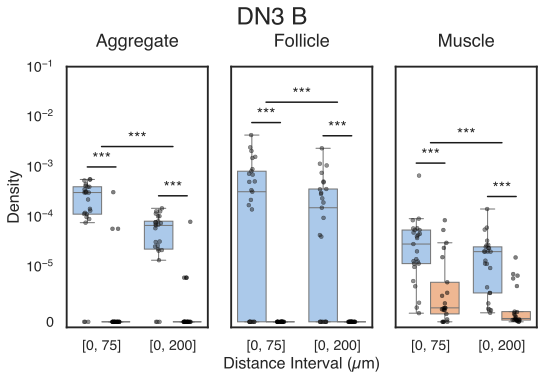

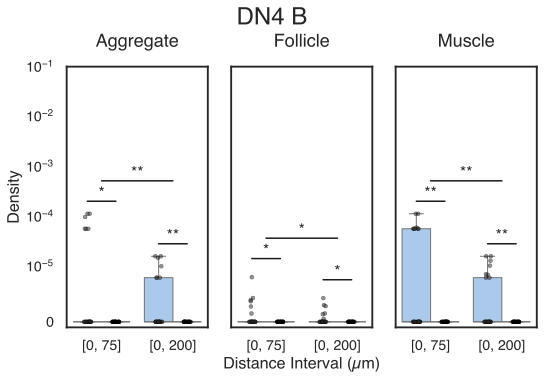

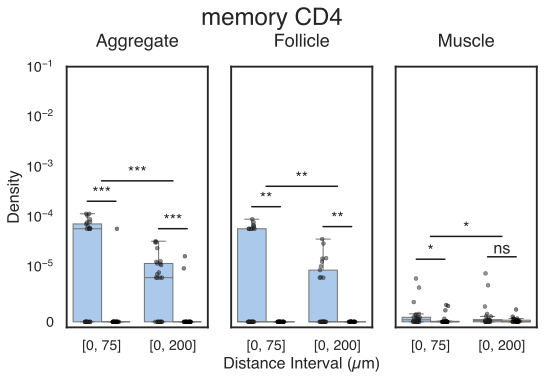

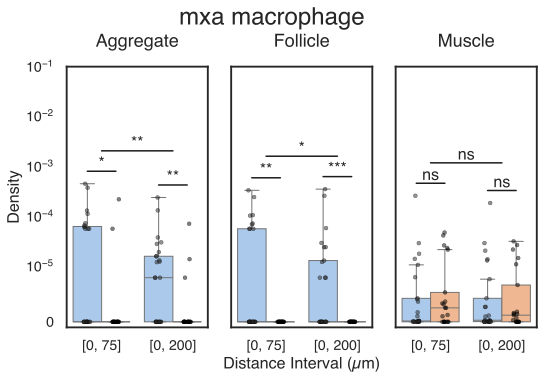

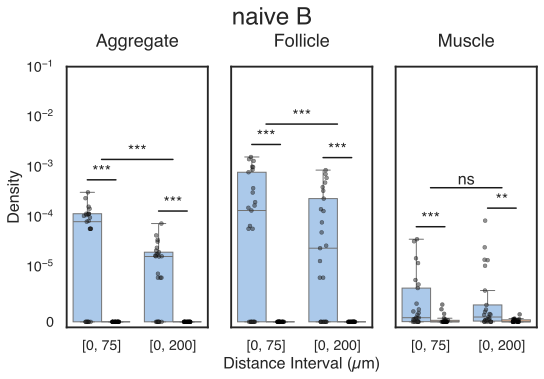

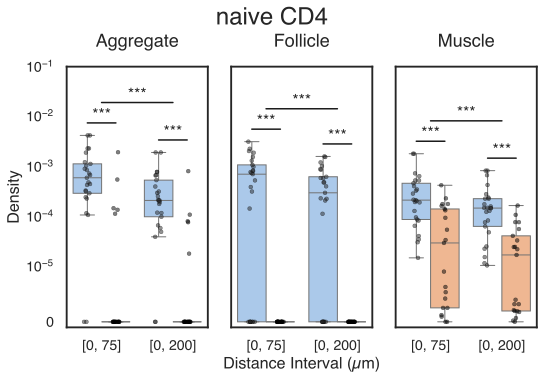

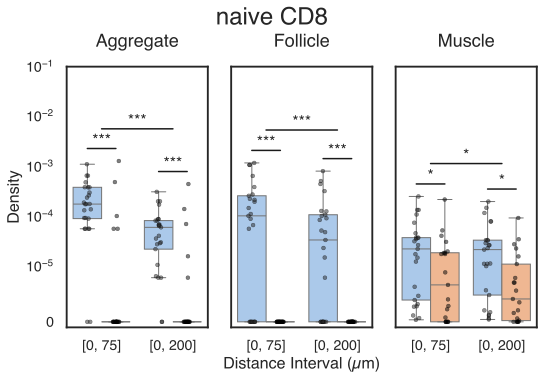

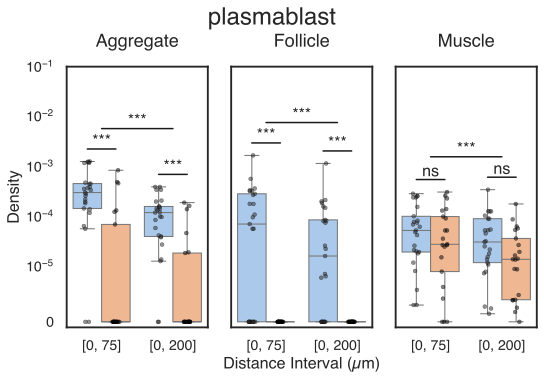

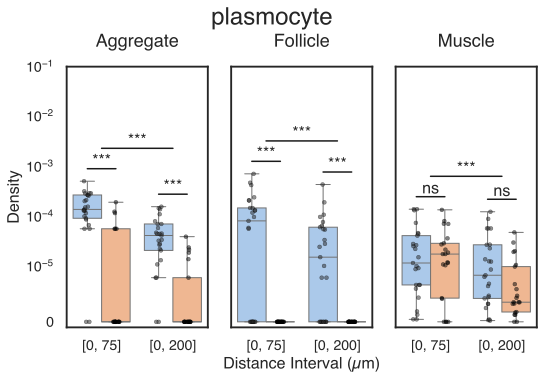

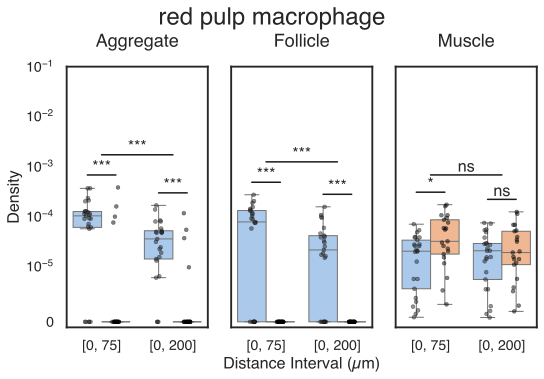

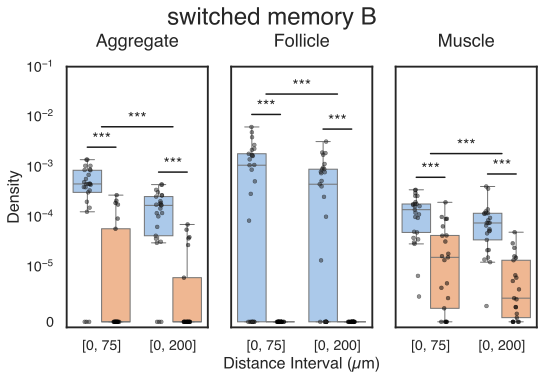

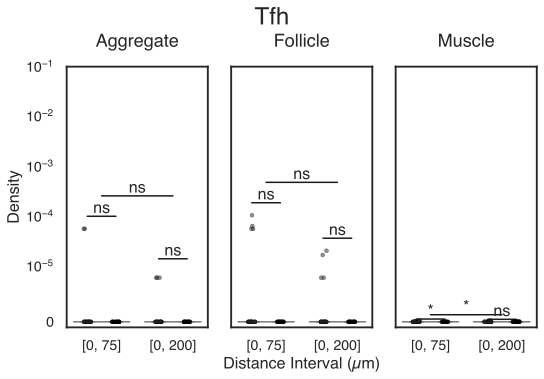

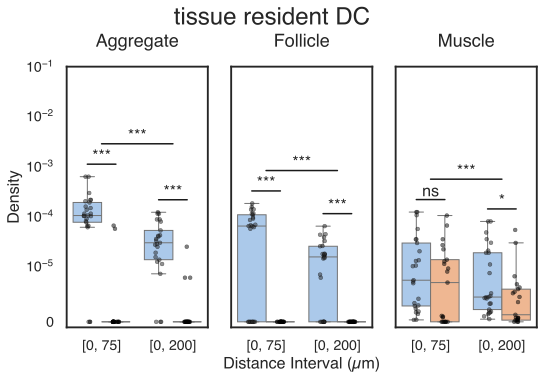

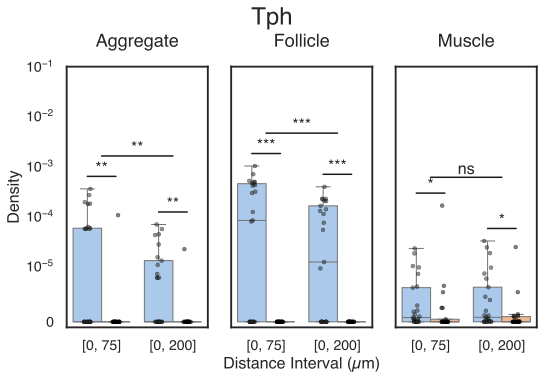

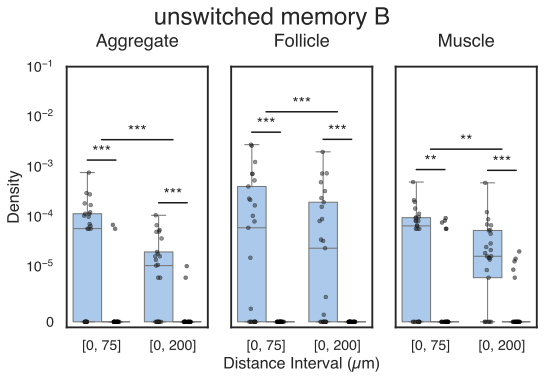

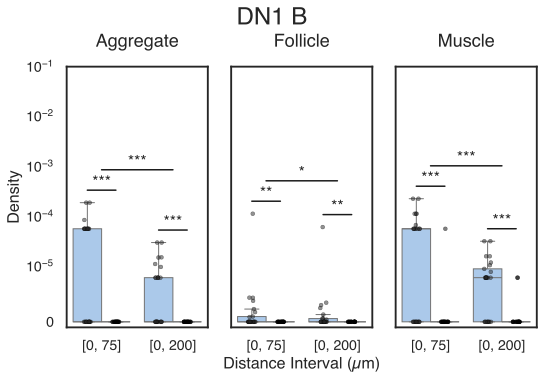

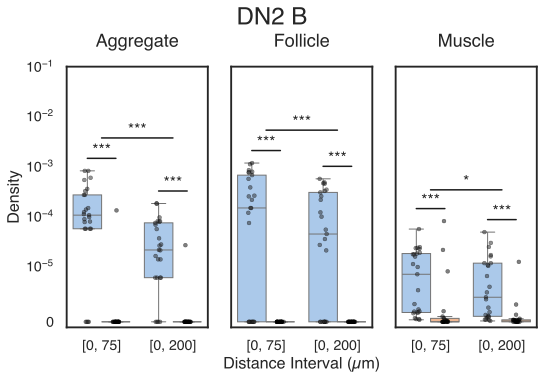

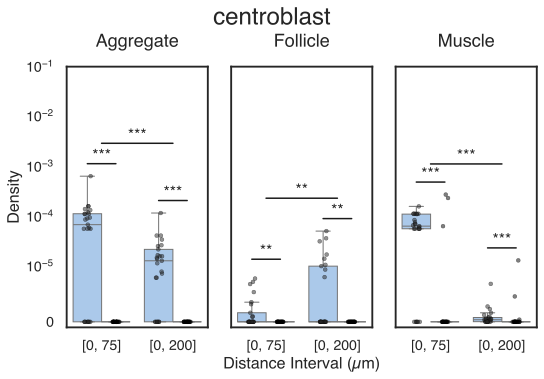

**Supplementary Figure 4:** Distance to b lymphocyte based enrichment of different immune cell types between BCM and non-BCM muscle tissue samples for each type of structure. (* : <0.05, ** : <0.01, *** : <0.001, ns: non-significant)

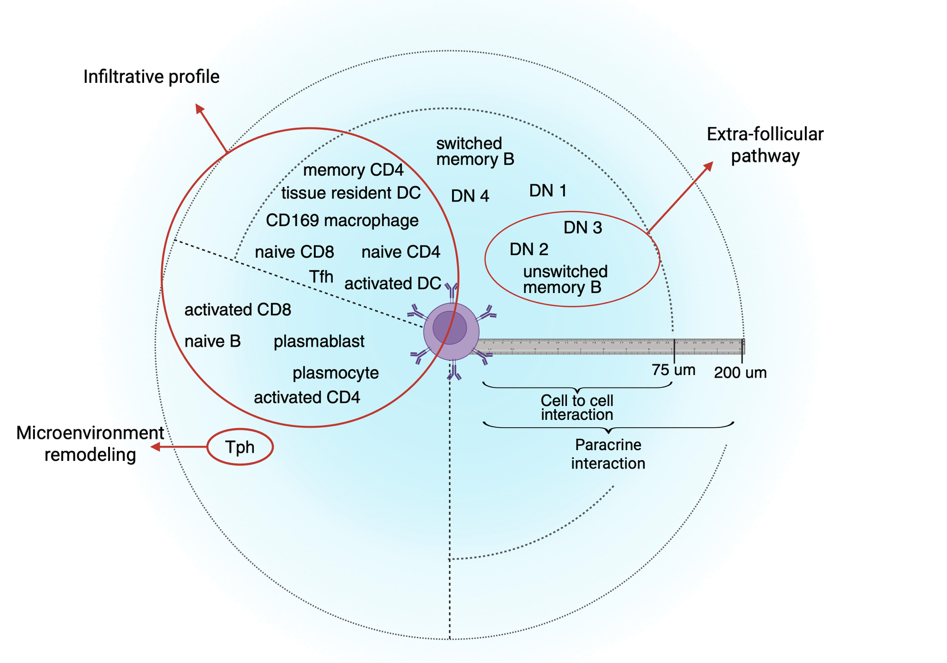

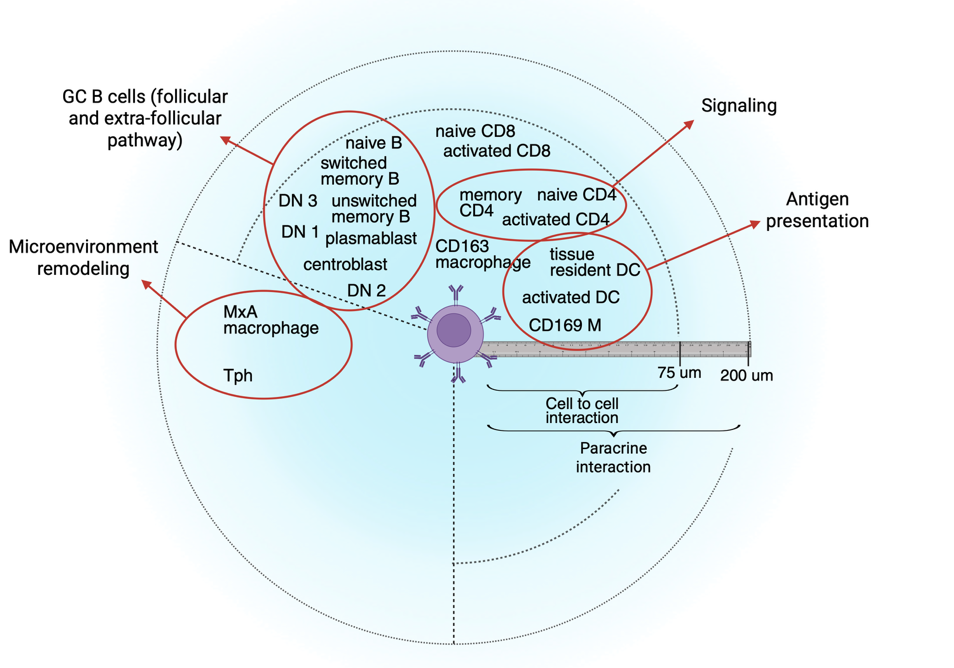

Follicle

Aggregate

Muscle

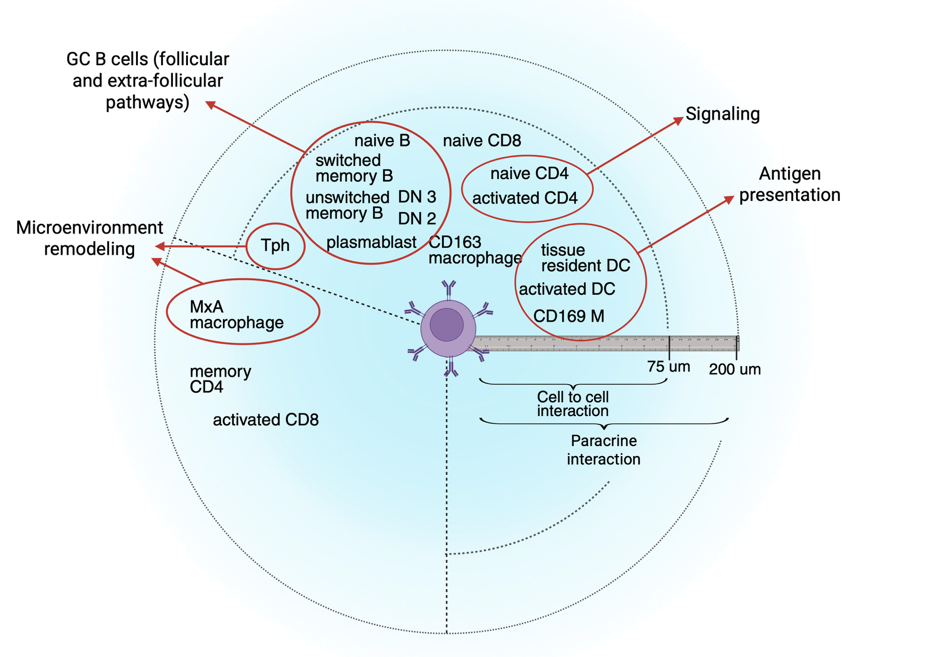

**Supplementary Figure 5:** Structure wise and distance to b lymphocyte-based enrichment of different immune cell types between BCM and non-BCM muscle tissue samples. (created with biorender)

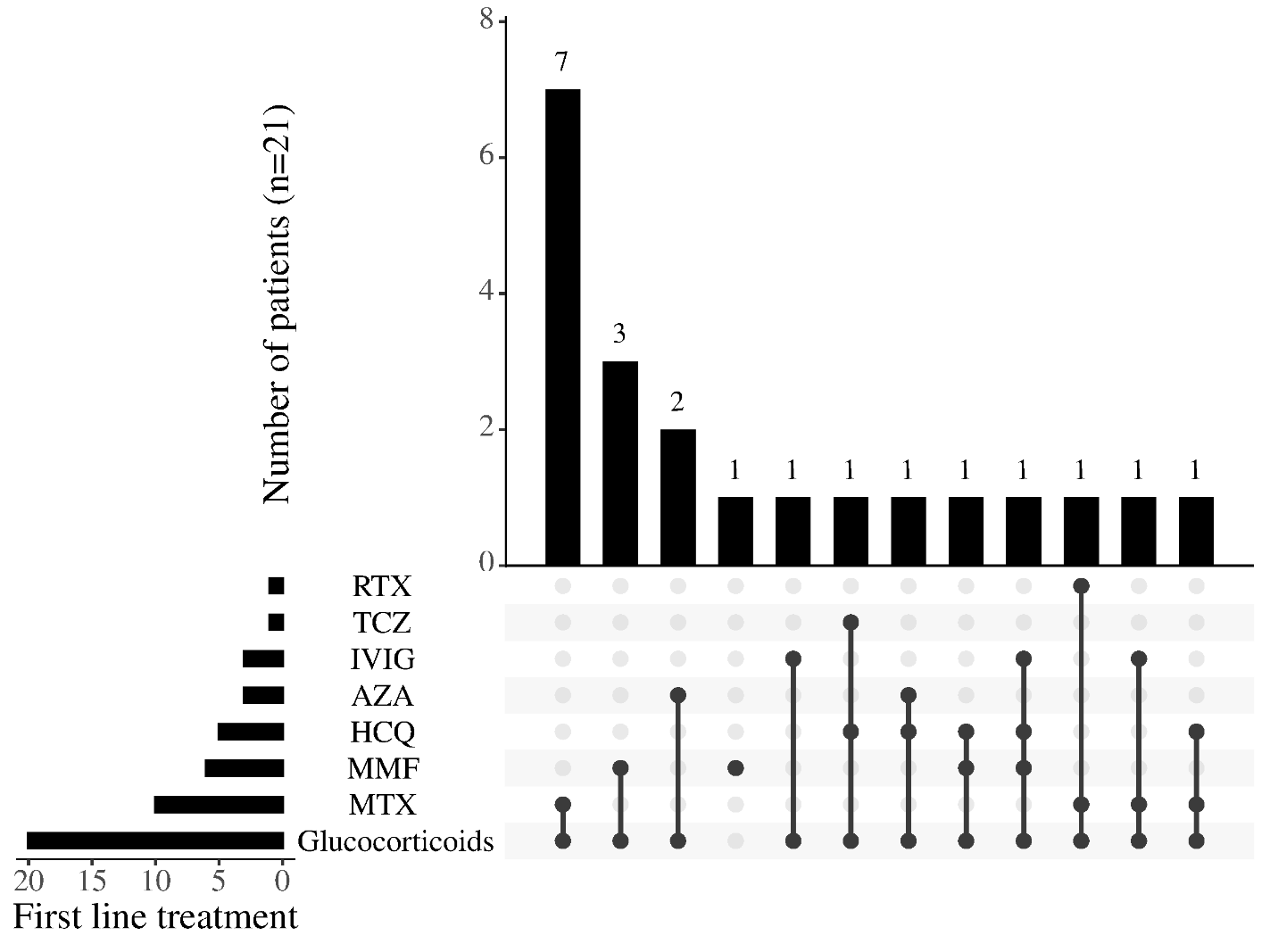

**Supplementary Figure 6**. First-line management of 21 patients with myositis with prominent B cell aggregates (BCM) with DMARDs.
RTX: rituximab, TCZ: tocilizumab, IVIG: intravenous immunoglobulin, AZA: azathioprine, HCQ: hydroxychloroquine, MMF: mycophenolate mofetil, MTX: methotrexate.
